# Low back pain service utilization and costs: association with timing of first-line services for individuals initially contacting a physician specialist. A retrospective cohort study

**DOI:** 10.1101/2023.03.22.23287530

**Authors:** David Elton, Meng Zhang

## Abstract

**Background:** Physician specialists (PS) are often the type of healthcare provider initially contacted by an individual with low back pain (LBP). LBP clinical practice guidelines (CPG) recommend a stepped approach to management with an emphasis on first-line non-pharmaceutical and non-interventional services.

**Objective:** Examine the association between the incorporation of CPG recommended first-line services, exposure to second- and third-line services and total episode cost for individuals with non-surgical LBP initially contacting a PS.

**Design:** Retrospective observational study with identical design to previous study focused on primary care physicians.

**Setting/Patients:** National sample of individuals with non-surgical LBP occurring in 2017-2019.

**Measurements:** Independent variables were initial contact with a PS, and the timing of incorporation of five types of first-line services. Dependent measures included exposure to thirteen types of health care services and total episode cost.

**Results:** 91,096 individuals were associated with 98,992 episodes of non-surgical LBP. 36.2% of the 33,277 PS initially contacted for an episode of LBP incorporated any first-line service at any time during an episode. A first-line service was provided in 24.0% of episodes with active care (19.5% of episodes), manual therapy (13.7%) and chiropractic manipulative therapy (6.5%) the most common. 7.3% of non-surgical LBP episodes included a first-line service within seven days of initial contact with a PS. These episodes were associated with a reduction in the use of prescription skeletal muscle relaxants (risk ratio (RR) 0.88) and opioids (RR 0.55), spinal injections (RR 0.84), and CT scans (RR 0.71), with no impact on the use of prescription NSAIDs, radiography, or MRI scans. First-line services were associated with an increase in total episode cost at any time of incorporation with chiropractic manipulation associated with the lowest cost increase. Younger individuals from zip codes with higher adjusted gross income were more likely to receive a first-line service in the first seven days of an episode.

**Limitations:** As a retrospective observational analysis of associations there are numerous potential confounders and limitations.

**Conclusions:** For individuals with non-surgical LBP PS provide second- or third-line services more frequently and earlier than CPG recommended first-line services. There is an opportunity to improve concordance with LBP CPGs for individuals with LBP initially contacting a PS.

## Introduction

The high prevalence and cost of low back pain (LBP) are well understood^1–4^ and considerable resources have been devoted to the development of high-quality LBP clinical practice guidelines (CPGs) that describe a stepped approach to management.^5–7^ Self-management, non-pharmacological and non-interventional services are recommended as first-line approaches for LBP without red flags of serious pathology.^5–7^ The timing of introduction of CPG recommended first-line services has been identified as being potentially important.^8–17^ Management of LBP that is not concordant with CPGs increases the risk of LBP transitioning from an acute to a chronic condition^18^ and is an important source of “low-value” care^19–22^, described as health care services generating cost without, or with minimal, beneficial impact on outcomes.^23,24^

The type of health care provider (HCP) initially contacted by an individual with LBP has been used as a method to evaluate variation in service utilization and cost outcomes.^25–27^ As specialists in the management of musculoskeletal conditions, it is not surprising that physician specialists (PS) like orthopedic surgeons, physical medicine and rehabilitation physicians, and pain management physicians are commonly consulted by individuals with LBP.^27^ Several barriers to PCP referral for CPG recommended first-line services have been identified.^28–33^ Compared to PCP management of LBP, the rate and timing of PS referral for first-line services is less well understood.

For individuals with non-surgical LBP initially contacting a PS, the aim of this retrospective, observational study was to examine the association between the timing of incorporation of active care (AC), manual therapy (MT), chiropractic manipulative therapy (CMT), osteopathic manipulative therapy (OMT), or acupuncture services, utilization of other healthcare services, and total cost. The hypothesis was physicians specializing in the management of LBP would be associated with a high degree of LBP CPG concordance and that early incorporation of one or more of these first-line services would be associated with lower rates of second- and third-line service use, and lower total episode cost.

## Methods

### Study design, population, setting and data sources

This is a retrospective cohort study of individuals initially contacting a PS for an episode of non-surgical LBP. The PS HCP category consisted of orthopedic surgeon, neurosurgeon, physical medicine and rehabilitation, pain management, neurology and rheumatology physician types.^27^ The study cohort was able to access all PS HCP types directly without a referral.

The study design was identical to a previous study involving primary care physicians (PCP).^34^ To facilitate a comparison between PCP and PS results, figure axis scales were kept constant. An enrollee database included de-identified enrollment records, and administrative claims data for individuals with LBP. De-identified HCP demographic information and professional licensure status was included in an HCP database. ZIP code level population race and ethnicity data was extracted from the US Census Bureau^35^, adjusted gross income (AGI) data from the Internal Revenue Service^36^ and socioeconomic status (SES) Area Deprivation Index (ADI) data, from the University of Wisconsin Neighborhood Atlas^®^ database.^37^ An analytic database was created by linking these multiple databases.

With study data de-identified or a Limited Data Set in compliance with the Health Insurance Portability and Accountability Act (HIPAA) and customer requirements the UnitedHealth Group Office of Human Research Affairs determined that this study was exempt from Institutional Review Board review. The study was conducted and reported based on the Strengthening the Reporting of Observational Studies in Epidemiology (STROBE) guidelines. (Supplement – STROBE Checklist).^38^

Like the identical PCP study^34^ the analysis was unable to control for numerous known, unknown, or unmeasurable confounders. In this analysis of associations, no attempt was made to compare results with the identical PCP study^34^, or to generate causal insights using potentially inadequate approaches such as propensity score matching^39^ to perform incomplete adjustment for typical confounders such as age, sex and non-LBP related co-morbidities.^40,41^ As an alternative, actual individual demographic and episodic characteristics and associations are reported for the timing of introduction of the first-line services analyzed in the study.

### Cohort selection and unit of analysis

The cohort consisted of individuals with continuous medical and pharmacy insurance coverage during the entire study period who were aged 18 years and older with a complete episode of LBP commencing and ending during 1/1/2017 to 12/31/2019. This timeframe was selected to follow the release of the American College of Physicians (ACP) LBP CPG^5^ in 2017 and before the influence of the COVID-19 epidemic on care patterns in early 2020.

Episode of care was selected as the unit of analysis. Episodes have been shown to be a valid way to organize administrative claims data associated with a condition.^42^ The *Symmetry* ® *Episode Treatment Groups*® *(ETG* ®) and *Episode Risk Groups*® *(ERG* ®) version 9.5 methodologies and definitions were used to translate administrative claims data into episodes, which have been reported as a valid measurement for comparison of HCPs based on cost of care.^43^ A previous study found a low risk of misclassification bias associated with using episode of care as the unit of analysis.^27^

For this study complete episodes were defined as having at least 91-day pre- and 61-day post-episode clean periods during which no services were provided by any HCP for any LBP diagnosis. Excluded from the analysis were LBP episodes including a surgical procedure, or associated with diagnoses of malignant and non-malignant neoplasms, fractures and other spinal trauma, infection, congenital deformities and scoliosis, autoimmune disorders, osteoporosis, and advanced arthritis. These exclusions are particularly important for LBP episodes where a PS was initially contacted and were made to address a potential study limitation of individuals with more complex conditions confounding the analysis of timing of incorporation of first-line non-pharmacological and non-interventional services.

### Variables

Data preprocessing, table generation, and initial analyses were performed using Python (*Python Language Reference, Version 3*.*7*.*5*., n.d.). A goodness of fit analysis was conducted using D’Agostino’s K-squared test. Non-normally distributed data are reported using the median and interquartile range (IQR).

The primary independent variables were initial contact with a PS, and the timing of incorporation of AC, MT, CMT, OMT, or acupuncture services. For LBP, these are the most frequently provided first-line services recommended by CPGs and covered by commercial insurance.^27^ Passive therapies, like ultrasound or electrical stimulation, were excluded from the definition of first-line services used in the analysis. For episodes initially contacting a PS the timing of incorporation of AC, MT, CMT, OMT, or acupuncture services was based on the number of days after the initial visit with a PS when these services were first billed by any type of HCP.

The primary dependent variable was the rate and timing of use of thirteen types of health care services segmented into first-, second-, and third-line service categories based on the ACP LBP CPG as a primary source for the designation.^5^ Secondary dependent variables included the total cost of care for all reimbursed services provided by any HCP during an episode, the number of different HCP seen during an episode, and episode duration measured in days. Total episode cost included costs associated with all services provided for an episode of LBP, including those not specifically identified in the thirteen categories used in the analyses. Costs for services for which an insurance claim was not submitted were not available. The episode duration was the number of days between the first and last date of service for each episode.

Bivariate analyses were performed comparing episode attributes associated with timing of introduction of AC, MT, CMT, OMT, or acupuncture services. For the bivariate analyses the reference baseline was episodes that did not include a specific first-line service. Fisher’s Exact test (p value of .001) was used for comparing the percent of episodes including a service, and Mann Whitney U test (p value of .001) was used for measures reported using median and IQR.

Using identical methods, all LBP tables, figures, and supplement items were replicated for non-surgical neck pain (NP) episodes where a PS was the initial HCP contacted. NP data are included as supplemental items using the same name as the corresponding LBP tables and figures.

## Results

The sample included 91,096 individuals, with a median age of 49 (Q1 38, Q3 57), and 54.9% females. These individuals were associated with 98,992 complete non-surgical LBP episodes involving 33,277 unique PS. There were $127,500,028 in reimbursed health care expenditures with a median total cost per episode of $389 (Q1 $138, Q3 $1,263). The median pre-episode clean period was 571 days (Q1 342, Q3 832). The median number of days between sequential episodes was 227 (Q1 126, Q3 364). The median post-episode clean period was 440 days (Q1 287, Q3 718) (Table 1). Individuals were from all 50 States and some U.S. territories. (Supplement - State)

**Table 1.** Cohort characteristics - % or Median (Q1, Q3)

|  |  |
| --- | --- |
| # of Individuals | 91096 |
| # of Episodes | 98992 |
| # of Specialist health care providers (HCP) | 33277 |
| Total cost (\$) | 127500028 |
| **Individuals with low back pain** |  |
| % Female | 54.9% |
| Age | 49 (38, 57) |
| ERG® risk score | 2.2 (1.1, 4.0) |
| **Individual home address zip code population attributes** |  |
| % non-Hispanic White (NHW) | 69.9% (49.6%, 83.1%) |
| Area Deprivation Index (ADI) | 40 (23, 58) |
| Household Adjusted Gross Income (AGI) (\$) | 71316 (53183, 105870) |
| HCP per 1000 - DC | 0.23 (0.10, 0.43) |
| HCP per 1000 - PT | 0.19 (0.05, 0.44) |
| HCP per 1000 - LAc | 0.00 (0.00, 0.04) |
| **Episode attributes** |  |
| Total cost (\$) | 389 (138, 1263) |
| # of HCP Seen | 2 (1, 4) |
| Episode duration - days (d) | 51 (1, 182) |
| Clean period - before initial episode (d) | 571 (342, 832) |
| Clean period - between sequential episodes (d) | 227 (126, 364) |
| Clean period - after final episode (d) | 440 (287, 718) |

The majority of non-surgical LBP episodes (76.0%) did not include a first-line service at any time during an episode. For the 24.0% of episodes that included any first-line service at any time AC (19.5% of episodes), MT (13.7%) and CMT (6.5%) were most common. OMT (0.8%) and acupuncture (0.5%) were rarely provided. Individuals were more likely to receive radiography (41.3%), prescription NSAIDs (24.8%), MRI (22.2%), opioids (18.8%), skeletal muscle relaxants (16.2% of episodes), and spinal injections (14.0%) than all first-line services except AC. (Table 2)

**Table 2.** Non-surgical low back pain initially contacting specialist - episodic service use by number of days (d) into episode when first-line service first provided

|  |  |  |  |  |  |  |  |  |  |  |  |  |  |  |  |
| --- | --- | --- | --- | --- | --- | --- | --- | --- | --- | --- | --- | --- | --- | --- | --- |
|  | Episodes |  | First Line |  |  |  |  | Passive Therapy | Second Line |  |  |  | Thirs Line |  |  |
|  | Count | % | AC | MT | CMT | OMT | Acu |  | Rx - NSAID | Rx - MM Relaxant | Imaging - Radiography | Imaging - MRI | Rx- Opioid | Spinal Injection | Imaging- CT |
| Total | 98992 | 100.0% | 19.5% | 13.7% | 6.5% | 0.8% | 0.5% | 7.4% | 24.8% | 16.2% | 41.3% | 22.2% | 18.8% | 14.0% | 2.3% |
| Any First Line Service |  |  |  |  |  |  |  |  |  |  |  |  |  |  |  |
| No First Line - reference | 75237 | 76.0% | 0.0% | 0.0% | 0.0% | 0.0% | 0.0% | 0.2% | 23.3% | 15.9% | 38.4% | 19.6% | 19.3% | 13.5% | 2.2% |
| 0-7d | 7209 | 7.3% | 84.8% | 61.0% | 18.2% | 7.4% | 1.9% | 29.9% | 23.5% | 14.0% | 44.9% | 18.8% | 11.3% | 11.4% | 1.6% |
| 8-14d | 3252 | 3.3% | 90.0% | 63.7% | 18.1% | 1.0% | 1.3% | 28.6% | 28.8% | 15.7% | 61.2% | 27.1% | 12.5% | 11.6% | 1.2% |
| 15-28d | 3531 | 3.6% | 85.6% | 59.1% | 22.9% | 1.1% | 1.2% | 28.0% | 27.6% | 17.0% | 56.4% | 35.9% | 16.1% | 13.7% | 1.6% |
| 29-60d | 3803 | 3.8% | 79.8% | 55.2% | 31.0% | 1.5% | 2.1% | 29.8% | 30.5% | 17.2% | 50.3% | 40.1% | 19.3% | 17.9% | 2.9% |
| 61-90d | 1840 | 1.9% | 72.2% | 51.6% | 41.6% | 1.2% | 1.8% | 30.3% | 34.3% | 19.1% | 49.4% | 36.4% | 23.8% | 20.4% | 3.5% |
| >90d | 4120 | 4.2% | 69.4% | 47.7% | 43.8% | 2.1% | 2.8% | 34.6% | 38.8% | 23.4% | 46.1% | 35.2% | 27.3% | 24.4% | 4.7% |
| Active Care (AC) |  |  |  |  |  |  |  |  |  |  |  |  |  |  |  |
| Not Provided - reference | 79709 | 80.5% | 0.0% | 1.3% | 4.3% | 0.7% | 0.3% | 2.0% | 23.5% | 15.9% | 38.0% | 19.3% | 19.4% | 13.3% | 2.2% |
| 0-7d | 5794 | 5.9% | 100.0% | 67.1% | 10.8% | 2.1% | 1.2% | 30.2% | 25.0% | 14.0% | 49.0% | 20.3% | 11.1% | 11.6% | 1.7% |
| 8-14d | 2940 | 3.0% | 100.0% | 68.2% | 10.6% | 0.6% | 0.6% | 28.3% | 28.9% | 15.4% | 64.8% | 28.1% | 11.4% | 11.8% | 1.3% |
| 15-28d | 3069 | 3.1% | 100.0% | 65.7% | 12.4% | 0.7% | 0.7% | 27.4% | 27.8% | 16.9% | 59.9% | 39.1% | 14.8% | 14.0% | 1.6% |
| 29-60d | 3071 | 3.1% | 100.0% | 64.1% | 17.2% | 0.7% | 1.3% | 28.5% | 31.0% | 17.5% | 55.7% | 46.5% | 17.8% | 19.9% | 3.2% |
| 61-90d | 1370 | 1.4% | 100.0% | 62.4% | 25.0% | 0.9% | 1.4% | 30.2% | 35.9% | 21.0% | 54.7% | 43.7% | 23.6% | 24.4% | 4.2% |
| > 90d | 3039 | 3.1% | 100.0% | 59.9% | 28.8% | 1.5% | 1.6% | 34.8% | 40.6% | 25.5% | 50.6% | 42.5% | 27.9% | 28.2% | 5.3% |
| Manual Therapy (MT) |  |  |  |  |  |  |  |  |  |  |  |  |  |  |  |
| Not Provided - reference | 85421 | 86.3% | 7.9% | 0.0% | 5.2% | 0.8% | 0.3% | 3.2% | 23.9% | 16.0% | 39.1% | 20.2% | 19.3% | 13.4% | 2.2% |
| 0-7d | 3616 | 3.7% | 93.1% | 100.0% | 10.7% | 1.0% | 1.7% | 35.1% | 23.8% | 14.6% | 46.1% | 19.9% | 10.8% | 12.2% | 1.4% |
| 8-14d | 1968 | 2.0% | 96.1% | 100.0% | 9.0% | 0.8% | 1.2% | 30.0% | 28.3% | 14.8% | 63.3% | 27.2% | 10.8% | 12.1% | 1.7% |
| 15-28d | 2321 | 2.3% | 94.8% | 100.0% | 10.9% | 0.9% | 1.0% | 32.0% | 27.7% | 16.9% | 60.4% | 38.2% | 14.4% | 14.3% | 1.5% |
| 29-60d | 2346 | 2.4% | 93.2% | 100.0% | 15.1% | 0.5% | 1.9% | 31.5% | 31.8% | 16.6% | 56.7% | 46.2% | 16.2% | 19.9% | 2.6% |
| 61-90d | 1047 | 1.1% | 88.0% | 100.0% | 22.3% | 0.7% | 2.1% | 34.4% | 35.1% | 19.1% | 55.2% | 44.5% | 23.9% | 25.6% | 3.9% |
| > 90d | 2273 | 2.3% | 87.5% | 100.0% | 25.5% | 1.5% | 2.6% | 39.0% | 42.1% | 26.6% | 51.6% | 44.8% | 27.5% | 30.1% | 5.2% |
| Manipulation - Chiropractic (CMT) |  |  |  |  |  |  |  |  |  |  |  |  |  |  |  |
| Not Provided - reference | 92533 | 93.5% | 17.5% | 12.5% | 0.0% | 0.8% | 0.4% | 4.9% | 24.4% | 16.1% | 41.4% | 22.3% | 18.6% | 14.0% | 2.3% |
| 0-7d | 1035 | 1.0% | 50.1% | 29.8% | 100.0% | 0.7% | 1.3% | 42.3% | 23.5% | 16.2% | 36.5% | 15.6% | 15.6% | 12.7% | 1.2% |
| 8-14d | 500 | 0.5% | 47.0% | 31.0% | 100.0% | 1.0% | 1.6% | 43.2% | 28.2% | 20.2% | 36.8% | 18.0% | 19.0% | 9.6% | 1.0% |
| 15-28d | 755 | 0.8% | 43.3% | 29.0% | 100.0% | 0.7% | 1.3% | 41.3% | 26.8% | 17.2% | 38.8% | 21.1% | 20.4% | 13.1% | 1.1% |
| 29-60d | 1185 | 1.2% | 44.5% | 28.6% | 100.0% | 0.8% | 1.5% | 41.7% | 28.7% | 16.4% | 34.3% | 17.5% | 23.7% | 11.2% | 1.5% |
| 61-90d | 829 | 0.8% | 45.8% | 32.2% | 100.0% | 0.5% | 1.2% | 39.4% | 32.0% | 16.8% | 38.0% | 20.4% | 22.9% | 13.0% | 1.3% |
| > 90d | 2155 | 2.2% | 49.8% | 32.2% | 100.0% | 0.9% | 2.0% | 47.0% | 35.5% | 20.6% | 44.3% | 24.8% | 23.7% | 19.1% | 3.3% |
| Manipulation - Osteopathic (OMT) |  |  |  |  |  |  |  |  |  |  |  |  |  |  |  |
| Not Provided - reference | 98225 | 99.2% | 19.4% | 13.7% | 6.5% | 0.0% | 0.4% | 7.4% | 24.9% | 16.3% | 41.4% | 22.2% | 18.9% | 14.0% | 2.3% |
| 0-7d | 499 | 0.5% | 26.5% | 9.8% | 4.6% | 100.0% | 0.8% | 12.0% | 10.2% | 7.8% | 15.0% | 6.6% | 5.6% | 7.0% | 0.8% |
| 8-14d | 29 | 0.0% | 31.0% | 20.7% | 0.0% | 100.0% | 0.0% | 20.7% | 24.1% | 17.2% | 34.5% | 13.8% | 13.8% | 24.1% | 6.9% |
| 15-28d | 38 | 0.0% | 42.1% | 31.6% | 7.9% | 100.0% | 0.0% | 15.8% | 23.7% | 23.7% | 28.9% | 18.4% | 23.7% | 15.8% | 2.6% |
| 29-60d | 57 | 0.1% | 35.1% | 22.8% | 8.8% | 100.0% | 5.3% | 19.3% | 15.8% | 19.3% | 31.6% | 22.8% | 24.6% | 26.3% | 3.5% |
| 61-90d | 25 | 0.0% | 28.0% | 24.0% | 4.0% | 100.0% | 0.0% | 20.0% | 24.0% | 24.0% | 36.0% | 20.0% | 16.0% | 32.0% | 4.0% |
| > 90d | 119 | 0.1% | 44.5% | 32.8% | 15.1% | 100.0% | 6.7% | 21.0% | 32.8% | 26.9% | 31.9% | 28.6% | 27.7% | 24.4% | 0.0% |
| Acupuncture (Acu) |  |  |  |  |  |  |  |  |  |  |  |  |  |  |  |
| Not Provided - reference | 98542 | 99.5% | 19.3% | 13.5% | 6.5% | 0.8% | 0.0% | 7.2% | 24.8% | 16.2% | 41.3% | 22.1% | 18.8% | 14.0% | 2.2% |
| 0-7d | 101 | 0.1% | 54.5% | 53.5% | 11.9% | 4.0% | 100.0% | 52.5% | 21.8% | 4.0% | 21.8% | 14.9% | 10.9% | 12.9% | 1.0% |
| 8-14d | 32 | 0.0% | 50.0% | 59.4% | 15.6% | 3.1% | 100.0% | 40.6% | 28.1% | 6.2% | 28.1% | 21.9% | 18.8% | 15.6% | 3.1% |
| 15-28d | 43 | 0.0% | 39.5% | 48.8% | 32.6% | 2.3% | 100.0% | 48.8% | 25.6% | 4.7% | 25.6% | 23.3% | 18.6% | 16.3% | 2.3% |
| 29-60d | 70 | 0.1% | 38.6% | 44.3% | 28.6% | 2.9% | 100.0% | 44.3% | 24.3% | 12.9% | 28.6% | 30.0% | 21.4% | 17.1% | 2.9% |
| 61-90d | 48 | 0.0% | 54.2% | 58.3% | 25.0% | 0.0% | 100.0% | 43.8% | 25.0% | 22.9% | 45.8% | 45.8% | 31.2% | 35.4% | 4.2% |
| > 90d | 156 | 0.2% | 48.1% | 52.6% | 25.6% | 4.5% | 100.0% | 53.2% | 38.5% | 16.7% | 34.0% | 32.7% | 21.8% | 26.3% | 4.5% |

Cells with red text denote that service usage was not significantly different from the referent of no first line service (Fishers's Exact p=>0.001)
Cells with black text denote that service usage was significantly different from the referent of no first line service (Fishers's Exact p=>0.001)
AC=Active Care, MT=Manual Therapy, CMT=Chiropractic Manipulative Treatment, OMT=Osteopathic Manipulative Treatment, Acu=Acupuncture

Within the first 7 days of an episode 7.3% of episodes included a first-line service with AC (5.9% of episodes), MT (3.7%) and CMT (1.0%) being most common. When introduced in the first seven days of an episode, AC was associated with a significant reduction in exposure to prescription opioids (risk ratio (RR) 0.57), CT scans (RR 0.76), spinal injections (RR 0.87), and skeletal muscle relaxants (RR 0.88). When introduced 8-14 days into an episode AC was associated with a significant reduction in exposure to prescription opioids (RR 0.58), CT scans (RR 0.58) and spinal injections (RR 0.88). At 15-28 days into an episode AC was associated with reductions in CT scans (RR 0.75) prescription opioids (RR 0.76). (Figure 1) MT was associated with similar reductions in exposure to prescription opioids. When introduced in the first seven days of an episode CMT was associated with significant reductions in CT scans (RR 0.51), MRI (RR 0.70), prescription opioids (RR 0.83), and radiography (RR 0.88). When introduced 8-14 days into an episode CMT was associated similar reductions in exposure to spinal injections (RR 0.69), and MRI (RR 0.81). (Figure 2) The small proportion of episodes including OMT or acupuncture at any time presented a barrier to identifying potential impact on second- and third-line service exposure. (Table 2) (Supplement – Risk Ratio)

**Figure 1.**
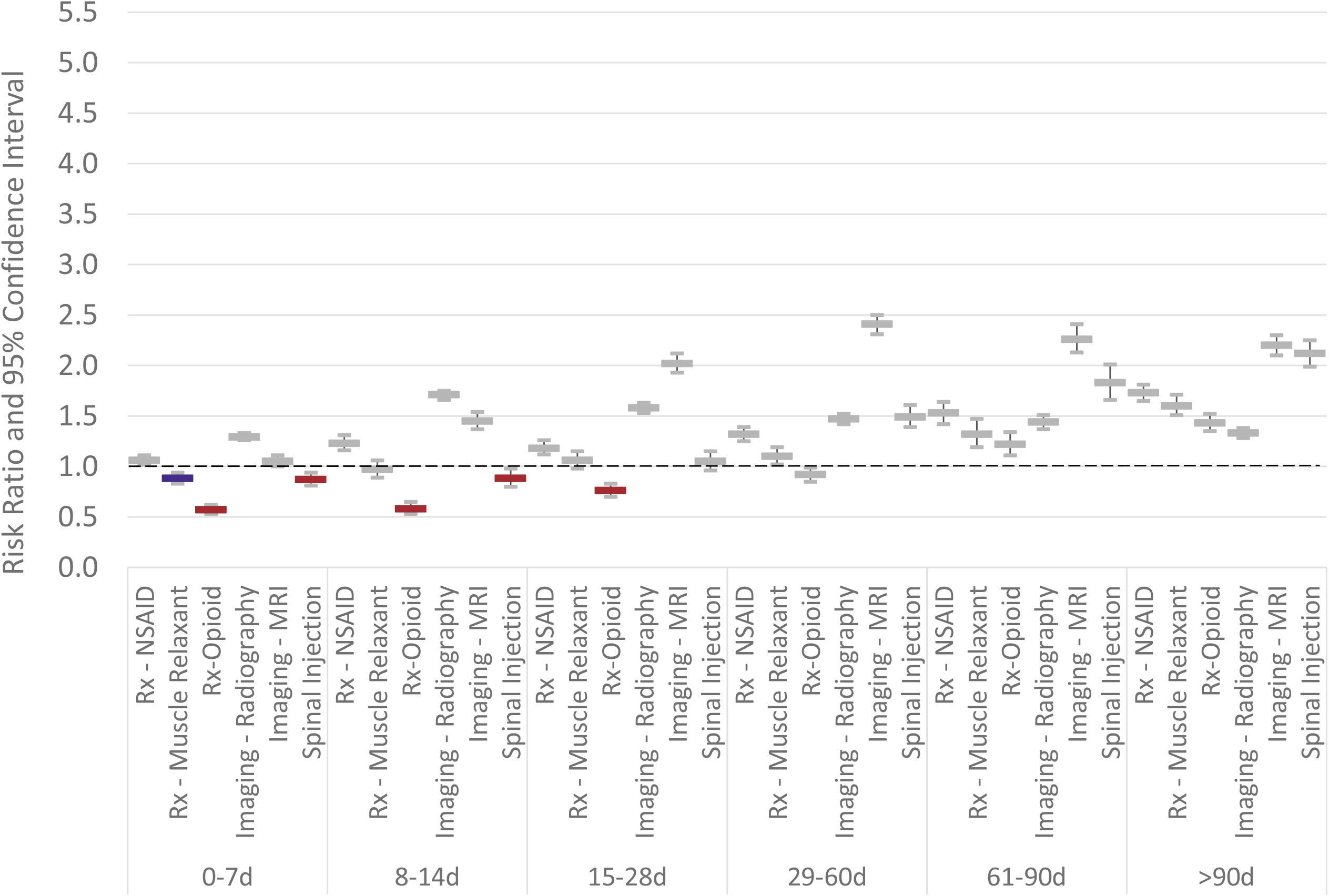
Individuals with non-surgical low back pain initially contacting a physician specialist. Risk ratio and 95% confidence interval for exposure to health care services based on timing of introduction of **active care** compared to episodes without active care

**Figure 2.**
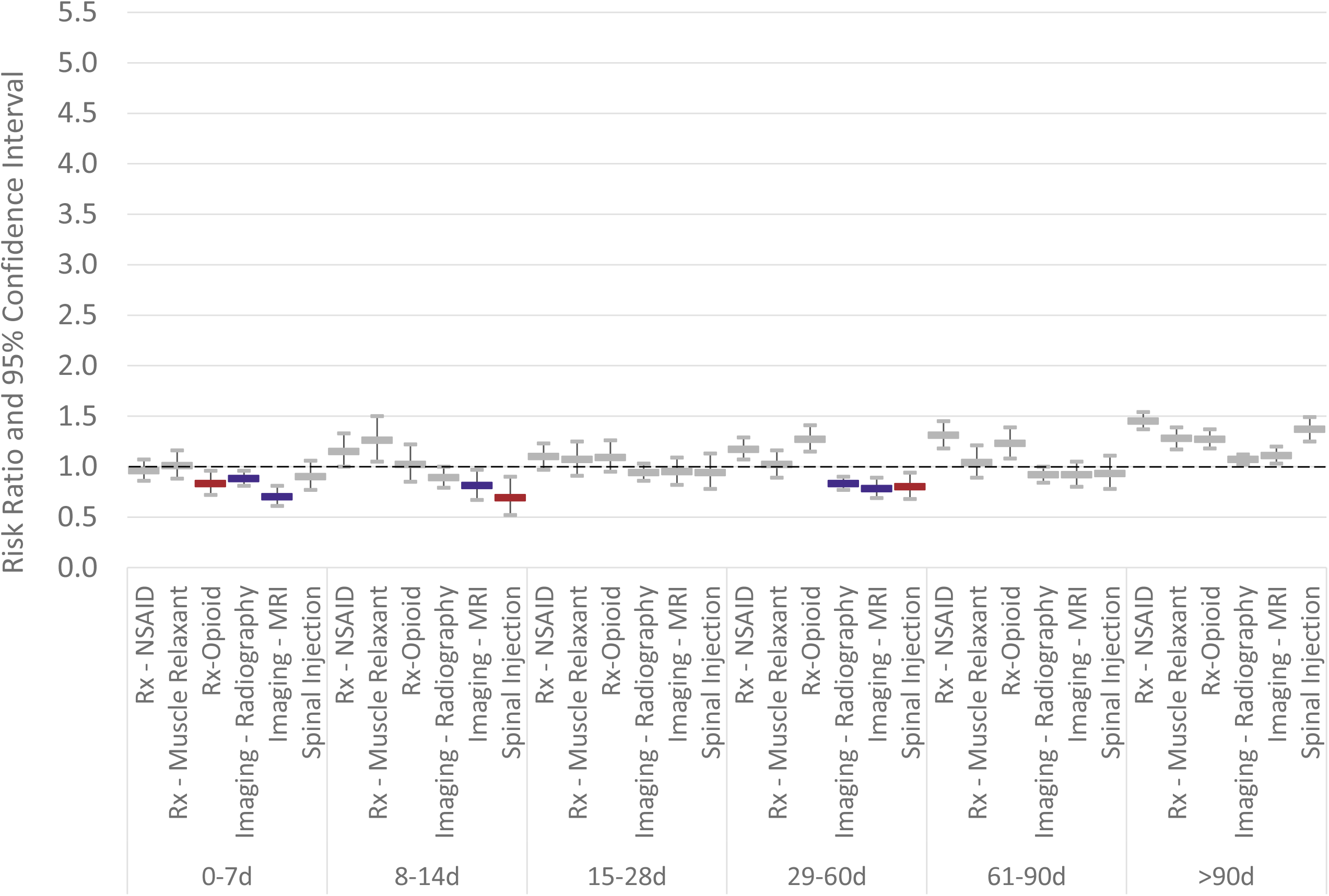
Individuals with non-surgical low back pain initially contacting a physician specialist. Risk ratio and 95% confidence interval for exposure to health care services based on timing of introduction of **chiropractic manipulation** compared to episodes without

For the 7.3% of episodes with a first-line service introduced in the first 7 days of an episode, and compared to episodes without a first-line service, individuals were younger (46 years old), with a lower *ERG*® risk score (1.7), from zip codes with lower deprivation (ADI 33), higher AGI (83,843), and with greater availability of a DC or PT. Among first-line services, acupuncture use was most strongly associated with lower deprivation (ADI 14), higher AGI (125,155), and greater availability of LAcs (0.05). (Figure 3) (Table 3)

**Table 3.**
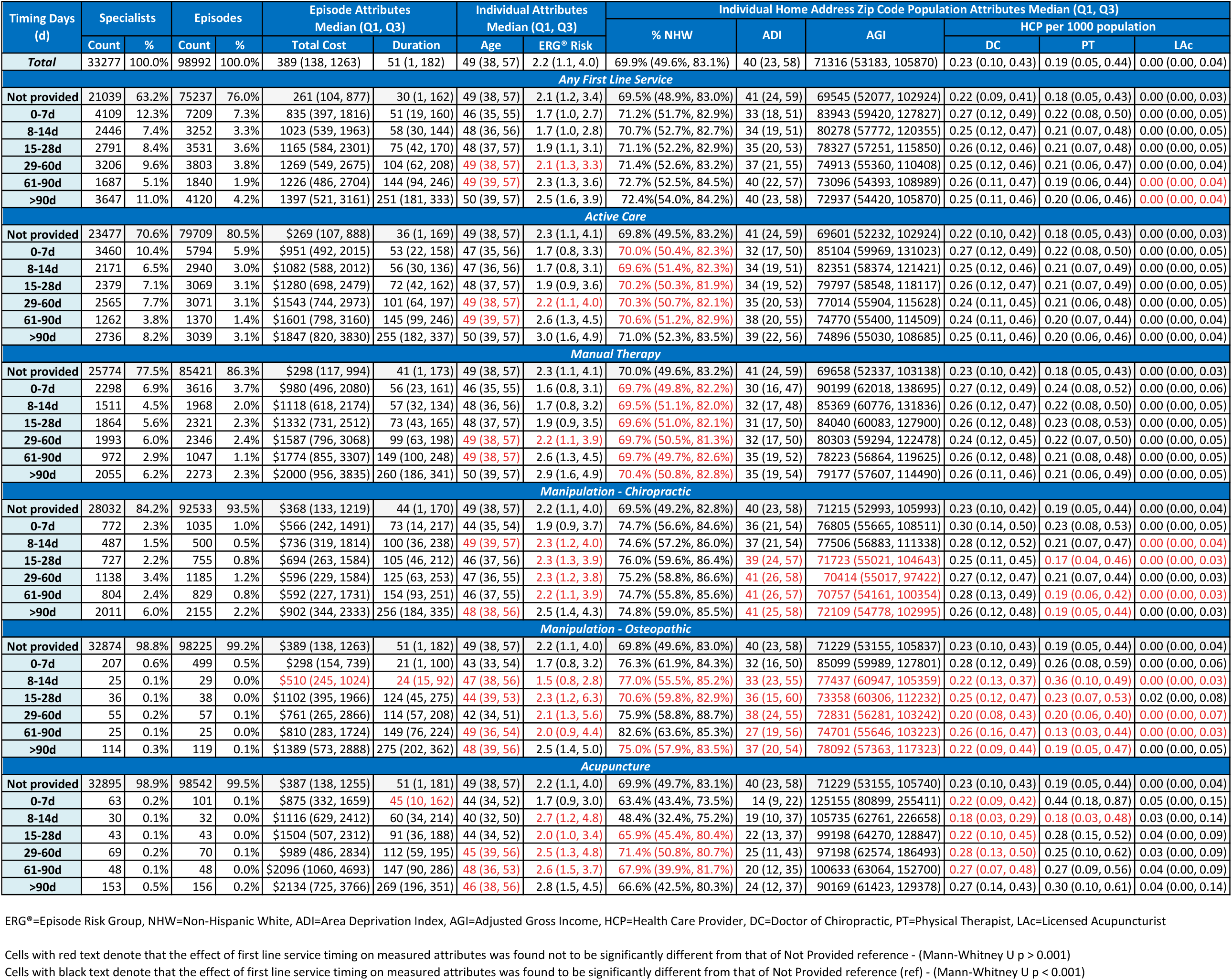
Individual, local population and episode attributes associated with individuals with low back pain initially contacting a specialist by timing of incorporation of first line services

**Figure 3.**
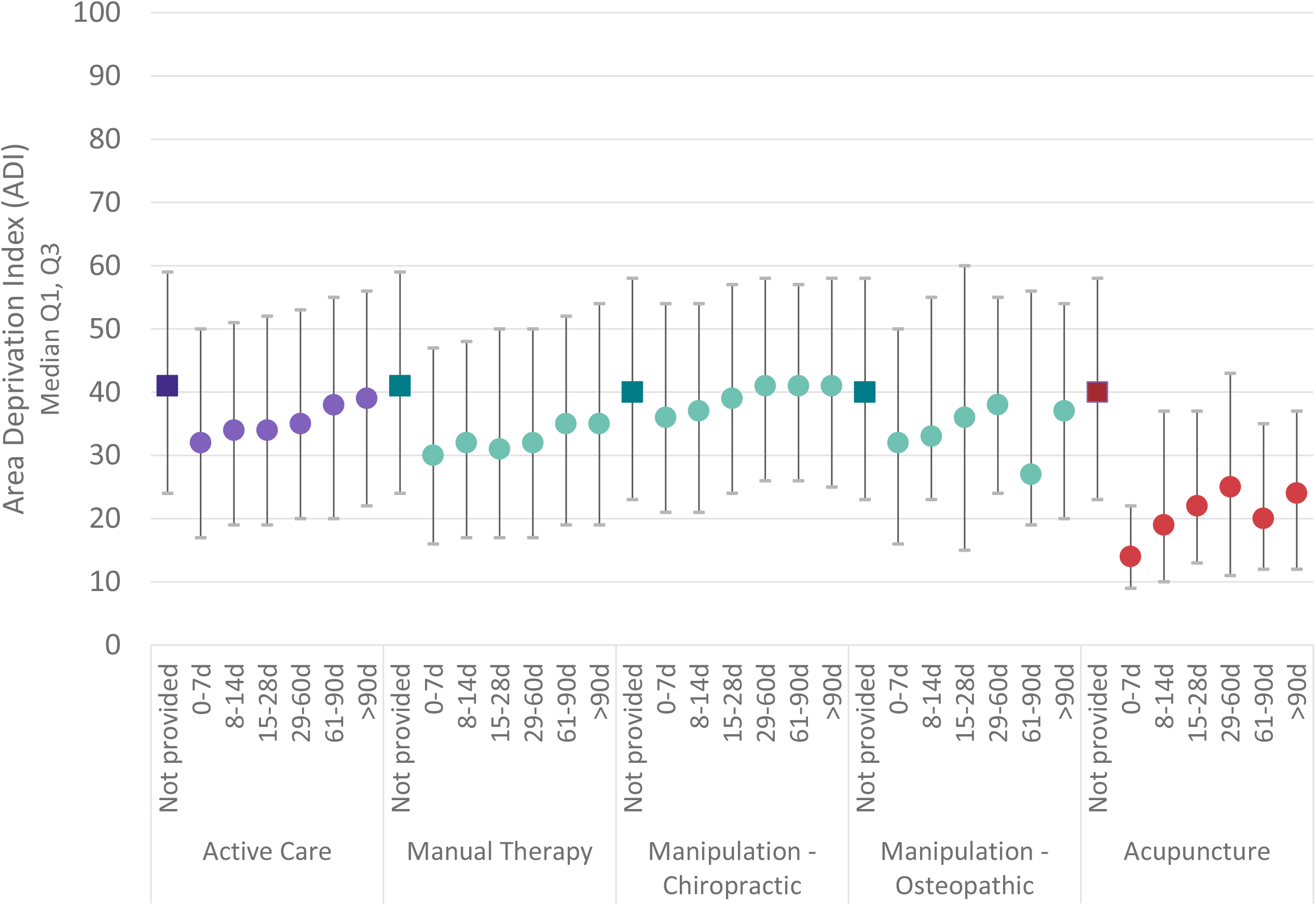
For individuals with low back pain initially contacting a physician specialist, Area Deprivation Index (ADI) of the individual’s home address zip code associated the number of days (d) into an episode when first line services are initially

Total episode cost increased with the introduction of any first-line service, and progressively increased the later a first line service was introduced. CMT and OMT were associated with the smallest total episode cost increase. (Figure 4) (Table 3)

**Figure 4.**
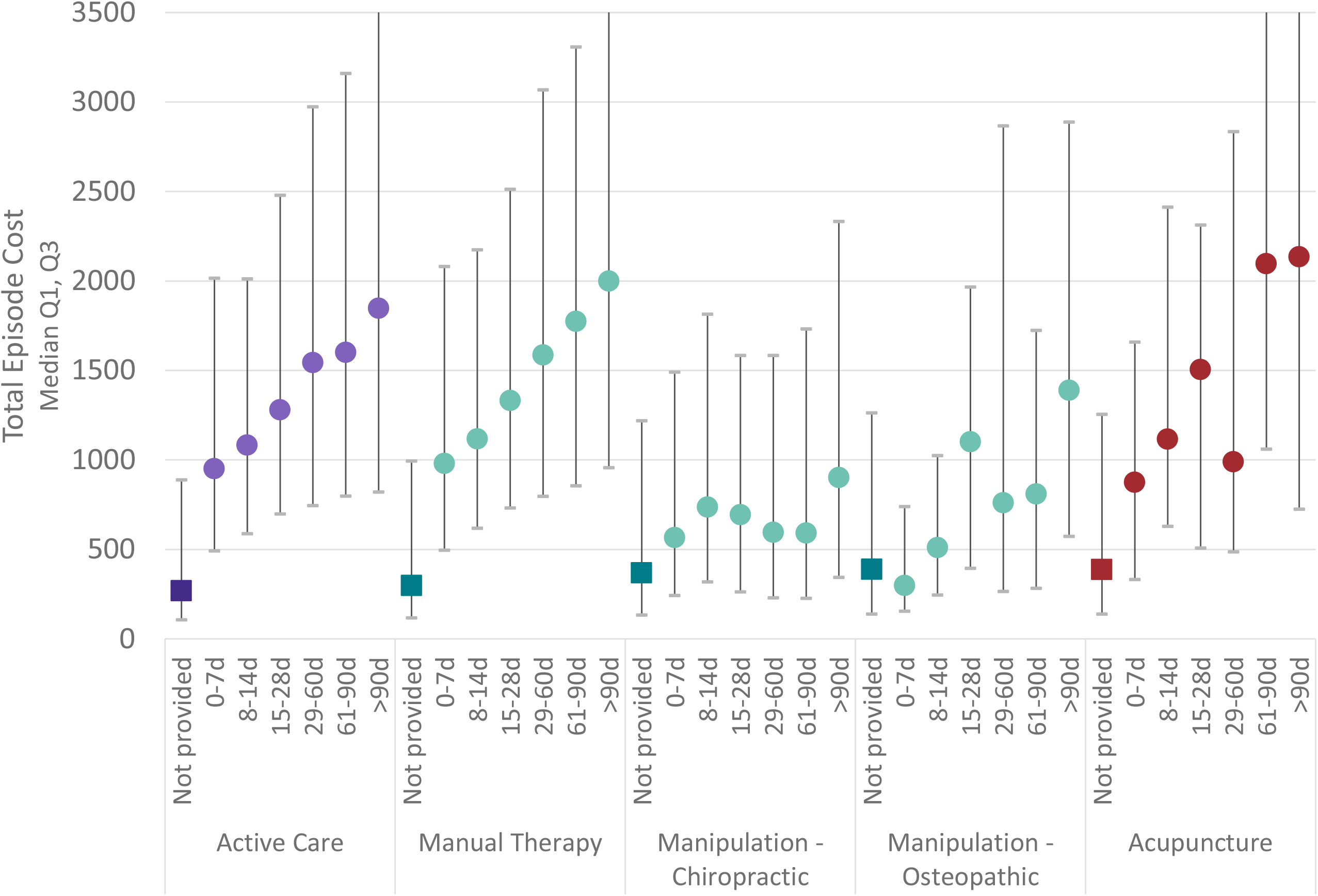
For individuals with low back pain initially contacting a physician specialist, total episode cost associated with number of days (d) into an episode when first line services are initially introduced

A statistical comparison of PS management of LBP and NP was not a study aim. Nevertheless, NP results are provided as supplemental items with results similar to LBP. LBP and NP cohort attributes were nearly identical except for cohort percent female where LBP was 54.9% and NP was 60.4%. (Table 1)(Supplement Table 1) The percent of episodes including any first-line service was 24.0% for LBP and 24.3% for NP. The first-line services most commonly incorporated during an episode was also similar; AC (19.5% for LBP, 19.3% for NP), MT (13.7%, 15.7%), CMT (6.5%, 6.7%), OMT (0.8%, 1.1%), and acupuncture (0.5%, 0.6%). (Table 2)(Supplement Table 2) Among first-line services CMT and OMT were associated the lowest total episode cost increase for both LBP and NP. (Table 3)(Supplement Table 3)

## Discussion

As specialists in the management of musculoskeletal conditions it was hypothesized that PS like orthopedic surgeons, physical medicine and rehabilitation, and pain management physicians, when initially contacted by individuals with LBP, would be associated with a high degree of CPG concordance. In contrast, this study found that PS incorporate CPG recommended non-pharmaceutical and non-interventional first-line services at any time in less than 25% of non-surgical LBP episodes, and that less than 15% of episodes include a first-line service in the first 14 days of an episode when the potential benefits of avoiding low-value second- and third-line services is highest. PS incorporate second- and third-line services more often than first-line services. These findings are nearly identical to an earlier study of PCP management of non-surgical LBP.^27^

As an observational retrospective cohort study of associations there are several limitations to consider. Perhaps most importantly a risk of selection bias is present due to the limited ability to control for individual preference for type of initial contact HCP, individual expectations or requests for specific health care services, and potentially meaningful differences in clinical complexity of individuals seeking treatment. These are particularly important potential limitations for individuals with LBP choosing to seek initial treatment from a PS. Following initial contact with a PS, and if not provided directly by the PS, it was not possible to differentiate whether first-line services were accessed by referral from the PS, by referral from another HCP, or accessed directly by the individual with LBP. These limitations were partially addressed by narrowing the study population by excluding LBP episodes associated with significant pathology and by focusing on only non-surgical LBP episodes.

While the cohort had continuous highly uniform commercial insurance coverage with quality and actuarial control measures applied to the processing of administrative claims data, data errors, variability in benefit plan design, variability in enrollee cost-sharing responsibility, and missing information were potential sources of confounding or bias. Even with the insurer HCP database being under continual validation it may have included errors or missing information regarding the identification of PS. Summarizing total episode cost has potential limitations associated with insurance coverage, nature of network participation, and alternative reimbursement models. While individuals from all 50 states and most US territories were included, providing a measure of generalizability, the cohort did not describe a U.S representative sample.

This study builds on and corroborates two earlier studies of the same underlying data. A low proportion of LBP episodes generally include timely incorporation of CPG recommended non-pharmacologic and non-interventional services.^27^ When initially contacted by an individual with LBP, PS are nearly identical to PCPs^34^ in that CPG recommended first-line services are infrequently incorporated, and if incorporated are typically later in an episode and in addition to second- and third-line services.

Administrative burden and the cost of CPG recommended first-line services have been identified as referral barriers.^30,31^ Like the previous identical PCP study^34^, this study found PS incorporate CPG recommended first-line services infrequently, and if incorporated it is after second- and third-line services, and resulted in higher total episode cost. The low rate of PS incorporation of CPG recommended first-line services may also reflect previously identified confounders like the time commitment to attend a visit, wait times to schedule a visit, transportation barriers, and a variety of individual characteristics.^29,44^

This study of PS management of LBP corroborates and expands on previous studies of PCP management of LBP that found earlier use of first-line services may be associated with a reduction in use of low value services and prescription pharmaceuticals, including opioids. ^13,14,34,45^ The study finding that the benefits of early use of first-line services are most evident if initiated within seven days of contacting a PS corroborates previous similar findings, ^11,34^ reinforces the principle that both the proportion of LBP episodes including, and timing of incorporation of, first-line services is important.

## Conclusion

High quality LBP CPGs recommend a stepped approach in which favorable natural history, self-care, and non-pharmacological services are emphasized as first-line approaches for individuals without red flags of serious pathology. For individuals with non-surgical LBP initially contacting a PS this study reveals second- and third-line pharmaceutical, imaging, and interventional services are provided more commonly and earlier than CPG recommended first-line services. With PS being positioned in the delivery system as musculoskeletal and spine specialists, identifying and addressing root causes of the observed high rate of CPG discordant LBP care is important.

## Supporting information

Supplement - Neck Pain - Figure 1

Supplement - Neck Pain - Figure 2

Supplement - Neck Pain - Figure 3

Supplement - Neck Pain - Figure 4

Supplement - Neck Pain - Risk Ratio

Supplement - Neck Pain - State Summary

Supplement - Neck Pain - Table 1

Supplement - Neck Pain - Table 2

Supplement - Neck Pain - Table 3

Supplement - Risk Ratio

Supplement - State Summary

Supplement - STROBE Checklist

## Data Availability

All data produced in the present work are contained in the manuscript

## Declarations

### Ethics approval and consent to participate

Because the data was de-identified or a Limited Data Set in compliance with the Health Insurance Portability and Accountability Act and customer requirements, the UnitedHealth Group Office of Human Research Affairs determined that this study was exempt from Institutional Review Board review.

### Consent for publication

Not applicable

### Availability of data and materials

The data are proprietary and are not available for public use but, under certain conditions, may be made available to editors and their approved auditors under a data-use agreement to confirm the findings of the current study.

### Authors’ contributions

Study conception and design; **DE.** Data acquisition; **DE, MZ.** Data analysis and interpretation; **DE, MZ.** Draft or substantially revise manuscript; **DE.**

## Acknowledgements

None

## List of Abbreviations

LBP: Low back pain
US: United States
CPG: Clinical practice guideline
PCP: Primary care provider
PS: Physician specialist
DC: Doctor of Chiropractic
PT: Physical Therapist
HCP: Health care provider
IQR: Interquartile range
AGI: Adjusted Gross Income
ADI: Area Deprivation Index
STROBE: Strengthening the Reporting of Observational Studies in Epidemiology
*ETG®*: Episode Treatment Group®
*ERG®*: Episode Risk Group®
ACP: American College of Physicians
CMT: Chiropractic manipulative treatment
OMT: Osteopathic manipulative treatment
AC: Active care
MT: Manual therapy

## References

1. Hartvigsen J, Hancock MJ, Kongsted A, et al. What low back pain is and why we need to pay attention. Lancet. Jun 9 2018;391(10137):2356–2367. doi:10.1016/S0140-6736(18)30480-X

2. Global burden of 369 diseases and injuries in 204 countries and territories, 1990-2019: a systematic analysis for the Global Burden of Disease Study 2019. Lancet. Oct 17 2020;396(10258):1204–1222. doi:10.1016/S0140-6736(20)30925-9

3. Buchbinder R, van Tulder M, Oberg B, et al. Low back pain: a call for action. Lancet. Jun 9 2018;391(10137):2384–2388. doi:10.1016/S0140-6736(18)30488-4

4. Dieleman JL, Cao J, Chapin A, et al. US Health Care Spending by Payer and Health Condition, 1996-2016. JAMA. Mar 3 2020;323(9):863–884. doi:10.1001/jama.2020.0734

5. Qaseem A, Wilt TJ, McLean RM, et al. Noninvasive Treatments for Acute, Subacute, and Chronic Low Back Pain: A Clinical Practice Guideline From the American College of Physicians. Ann Intern Med. Apr 4 2017;166(7):514–530. doi:10.7326/M16-2367

6. Meroni R, Piscitelli D, Ravasio C, et al. Evidence for managing chronic low back pain in primary care: a review of recommendations from high-quality clinical practice guidelines. Disabil Rehabil. Apr 2021;43(7):1029–1043. doi:10.1080/09638288.2019.1645888

7. Foster NE, Anema JR, Cherkin D, et al. Prevention and treatment of low back pain: evidence, challenges, and promising directions. Lancet. Jun 9 2018;391(10137):2368–2383. doi:10.1016/S0140-6736(18)30489-6

8. Wirth B, Riner F, Peterson C, et al. An observational study on trajectories and outcomes of chronic low back pain patients referred from a spine surgery division for chiropractic treatment. Chiropr Man Therap. 2019;27:6. doi:10.1186/s12998-018-0225-8

9. Rundell SD, Gellhorn AC, Comstock BA, Heagerty PJ, Friedly JL, Jarvik JG. Clinical outcomes of early and later physical therapist services for older adults with back pain. Spine J. Aug 1 2015;15(8):1744–55. doi:10.1016/j.spinee.2015.04.001

10. Ojha HA, Wyrsta NJ, Davenport TE, Egan WE, Gellhorn AC. Timing of Physical Therapy Initiation for Nonsurgical Management of Musculoskeletal Disorders and Effects on Patient Outcomes: A Systematic Review. J Orthop Sports Phys Ther. Feb 2016;46(2):56–70. doi:10.2519/jospt.2016.6138

11. Liu X, Hanney WJ, Masaracchio M, et al. Immediate Physical Therapy Initiation in Patients With Acute Low Back Pain Is Associated With a Reduction in Downstream Health Care Utilization and Costs. Physical Therapy. 2018;98(5):336–347. doi:10.1093/ptj/pzy023

12. Halfpap J, Riebel L, Tognoni A, Coller M, Sheu RG, Rosenthal MD. Improving Access and Decreasing Healthcare Utilization for Patients With Acute Spine Pain: Five-Year Results of a Direct Access Clinic. Mil Med. Mar 12 2022;doi:10.1093/milmed/usac064

13. Fritz JM, Lane E, McFadden M, et al. Physical Therapy Referral From Primary Care for Acute Back Pain With Sciatica : A Randomized Controlled Trial. Ann Intern Med. Jan 2021;174(1):8–17. doi:10.7326/M20-4187

14. Fritz JM, Kim M, Magel JS, Asche CV. Cost-Effectiveness of Primary Care Management With or Without Early Physical Therapy for Acute Low Back Pain: Economic Evaluation of a Randomized Clinical Trial. Spine (Phila Pa 1976). Mar 2017;42(5):285–290. doi:10.1097/BRS.0000000000001729

15. Foster NE, Konstantinou K, Lewis M, et al. Stratified versus usual care for the management of primary care patients with sciatica: the SCOPiC RCT. Health Technol Assess. Oct 2020;24(49):1–130. doi:10.3310/hta24490

16. Elder C, DeBar L, Ritenbaugh C, et al. Comparative Effectiveness of Usual Care With or Without Chiropractic Care in Patients with Recurrent Musculoskeletal Back and Neck Pain. J Gen Intern Med. Sep 2018;33(9):1469–1477. doi:10.1007/s11606-018-4539-y

17. Bezdjian S, Whedon JM, Russell R, Goehl JM, Kazal LA, Jr. Efficiency of primary spine care as compared to conventional primary care: a retrospective observational study at an Academic Medical Center. Chiropr Man Therap. Jan 6 2022;30(1):1. doi:10.1186/s12998-022-00411-x

18. Stevans JM, Delitto A, Khoja SS, et al. Risk Factors Associated With Transition From Acute to Chronic Low Back Pain in US Patients Seeking Primary Care. JAMA Netw Open. Feb 1 2021;4(2):e2037371. doi:10.1001/jamanetworkopen.2020.37371

19. AlEissa SI, Tamai K, Konbaz F, et al. SPINE20 A global advocacy group promoting evidence-based spine care of value. Eur Spine J. Aug 2021;30(8):2091–2101. doi:10.1007/s00586-021-06890-5

20. Buchbinder R, Underwood M, Hartvigsen J, Maher CG. The Lancet Series call to action to reduce low value care for low back pain: an update. Pain. Sep 2020;161 Suppl 1:S57–S64. doi:10.1097/j.pain.0000000000001869

21. Cassel CK, Guest JA. Choosing wisely: helping physicians and patients make smart decisions about their care. JAMA. May 2 2012;307(17):1801–2. doi:10.1001/jama.2012.476

22. Colla CH, Morden NE, Sequist TD, Schpero WL, Rosenthal MB. Choosing wisely: prevalence and correlates of low-value health care services in the United States. J Gen Intern Med. Feb 2015;30(2):221–8. doi:10.1007/s11606-014-3070-z

23. Schwartz AL, Landon BE, Elshaug AG, Chernew ME, McWilliams JM. Measuring low-value care in Medicare. JAMA Intern Med. Jul 2014;174(7):1067–76. doi:10.1001/jamainternmed.2014.1541

24. Reid RO, Rabideau B, Sood N. Low-Value Health Care Services in a Commercially Insured Population. JAMA Intern Med. Oct 1 2016;176(10):1567–1571. doi:10.1001/jamainternmed.2016.5031

25. Fritz JM, Kim J, Dorius J. Importance of the type of provider seen to begin health care for a new episode low back pain: associations with future utilization and costs. Journal of Evaluation in Clinical Practice. 2016;22(2):247–252. doi:https://doi.org/10.1111/jep.12464

26. Kazis LE, Ameli O, Rothendler J, et al. Observational retrospective study of the association of initial healthcare provider for new-onset low back pain with early and long-term opioid use. BMJ Open. Sep 20 2019;9(9):e028633. doi:10.1136/bmjopen-2018-028633

27. Elton D, Kosloff TM, Zhang M, et al. Low back pain care pathways and costs: association with the type of initial contact health care provider. A retrospective cohort study (preprint). medRxiv. 2022:2022.06.17.22276443. doi:10.1101/2022.06.17.22276443

28. Zheng P, Kao MC, Karayannis NV, Smuck M. Stagnant Physical Therapy Referral Rates Alongside Rising Opioid Prescription Rates in Patients With Low Back Pain in the United States 1997-2010. Spine (Phila Pa 1976). May 1 2017;42(9):670–674. doi:10.1097/BRS.0000000000001875

29. Stevens GL. Behavioral and access barriers to seeking chiropractic care: a study of 3 New York clinics. J Manipulative Physiol Ther. Oct 2007;30(8):566–72. doi:10.1016/j.jmpt.2007.07.015

30. Roseen EJ, Conyers FG, Atlas SJ, Mehta DH. Initial Management of Acute and Chronic Low Back Pain: Responses from Brief Interviews of Primary Care Providers. J Altern Complement Med. Mar 2021;27(S1):S106–S114. doi:10.1089/acm.2020.0391

31. Penney LS, Ritenbaugh C, Elder C, Schneider J, Deyo RA, DeBar LL. Primary care physicians, acupuncture and chiropractic clinicians, and chronic pain patients: a qualitative analysis of communication and care coordination patterns. BMC Complement Altern Med. Jan 25 2016;16:30. doi:10.1186/s12906-016-1005-4

32. Greene BR, Smith M, Allareddy V, Haas M. Referral patterns and attitudes of primary care physicians towards chiropractors. BMC Complement Altern Med. Mar 1 2006;6:5. doi:10.1186/1472-6882-6-5

33. Coulter ID, Singh BB, Riley D, Der-Martirosian C. Interprofessional referral patterns in an integrated medical system. J Manipulative Physiol Ther. Mar-Apr 2005;28(3):170–4. doi:10.1016/j.jmpt.2005.02.016

34. Elton D, Zhang M. Low back pain service utilization and costs: association with timing of first-line services for individuals initially contacting a primary care provider. A retrospective cohort study. medRxiv. 2022:2022.06.30.22277102. doi:10.1101/2022.06.30.22277102

35. US Census Bureau - ACS Demographic and Housing Estimates. Accessed 12/14/2022. https://data.census.gov/table?q=United+States&g=0100000US&y=2020

36. Internal Revenue Service - US Department of Treasury SOI tax stats - individual income tax statistics - 2019 ZIP code data. Accessed 12/14/2022. https://www.irs.gov/statistics/soi-tax-stats-individual-income-tax-statistics-2019-zip-code-data-soi

37. Kind AJH, Buckingham WR. Making Neighborhood-Disadvantage Metrics Accessible - The Neighborhood Atlas. N Engl J Med. Jun 28 2018;378(26):2456–2458. doi:10.1056/NEJMp1802313

38. von Elm E, Altman DG, Egger M, et al. The Strengthening the Reporting of Observational Studies in Epidemiology (STROBE) Statement: guidelines for reporting observational studies. Int J Surg. Dec 2014;12(12):1495–9. doi:10.1016/j.ijsu.2014.07.013

39. King G NR. Why Propensity Scores Should Not Be Used for Matching. Political Analysis. 2019;27(4):435–454. doi:10.1017/pan.2019.11

40. Smith GD, Ebrahim S. Data dredging, bias, or confounding. BMJ. Dec 21 2002;325(7378):1437–8. doi:10.1136/bmj.325.7378.1437

41. Hernan MA. The C-Word: Scientific Euphemisms Do Not Improve Causal Inference From Observational Data. Am J Public Health. May 2018;108(5):616–619. doi:10.2105/AJPH.2018.304337

42. Peterson C, Grosse SD, Dunn A. A practical guide to episode groupers for cost-of-illness analysis in health services research. SAGE Open Med. 2019;7:2050312119840200. doi:10.1177/2050312119840200

43. Insight O. Symmetry episode treatment groups: measuring health care with meaningful episodes of care. 2012.

44. MacKay C, Hawker G, Jaglal S. A qualitative study exploring the barriers and facilitators to optimal physical therapy management of early knee osteoarthritis symptoms. Osteoarthritis and Cartilage. 2017;25:S396. doi:10.1016/j.joca.2017.02.681

45. Fritz JM, Childs JD, Wainner RS, Flynn TW. Primary care referral of patients with low back pain to physical therapy: impact on future health care utilization and costs. Spine (Phila Pa 1976). Dec 1 2012;37(25):2114–21. doi:10.1097/BRS.0b013e31825d32f5

