## Supplement - Neck Pain - Figure 1 for "Low back pain service utilization and costs: association with timing of first-line services for individuals initially contacting a physician specialist. A retrospective cohort study"

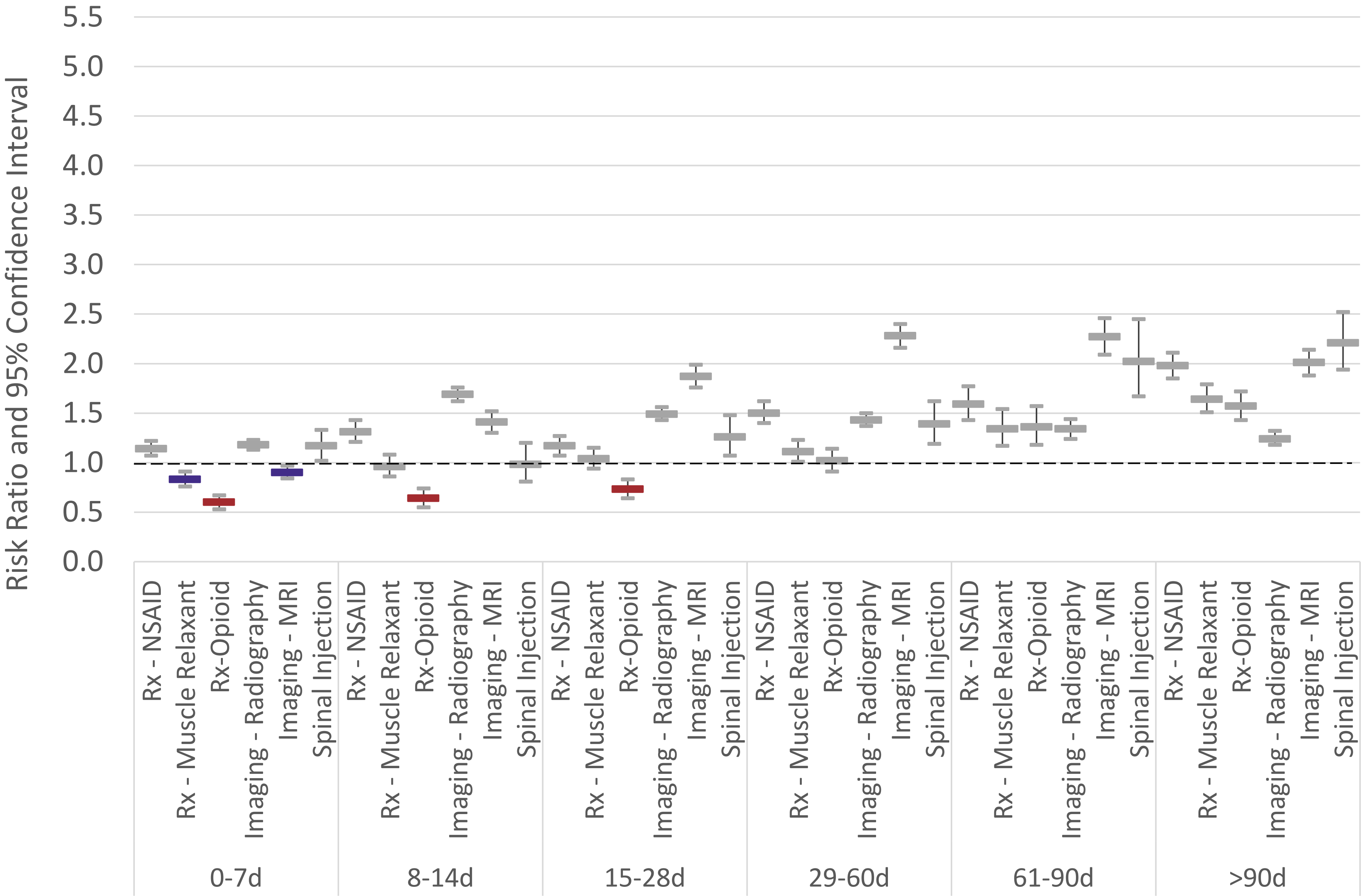


**Supplemental Figure 1.** Individuals with non-surgical neck pain initially contacting a physician specialist. Risk ratio and 95% confidence interval for exposure to health care services based on timing of introduction of **active care** compared to episodes without active care
