## Supplement - Neck Pain - Figure 2 for "Low back pain service utilization and costs: association with timing of first-line services for individuals initially contacting a physician specialist. A retrospective cohort study"

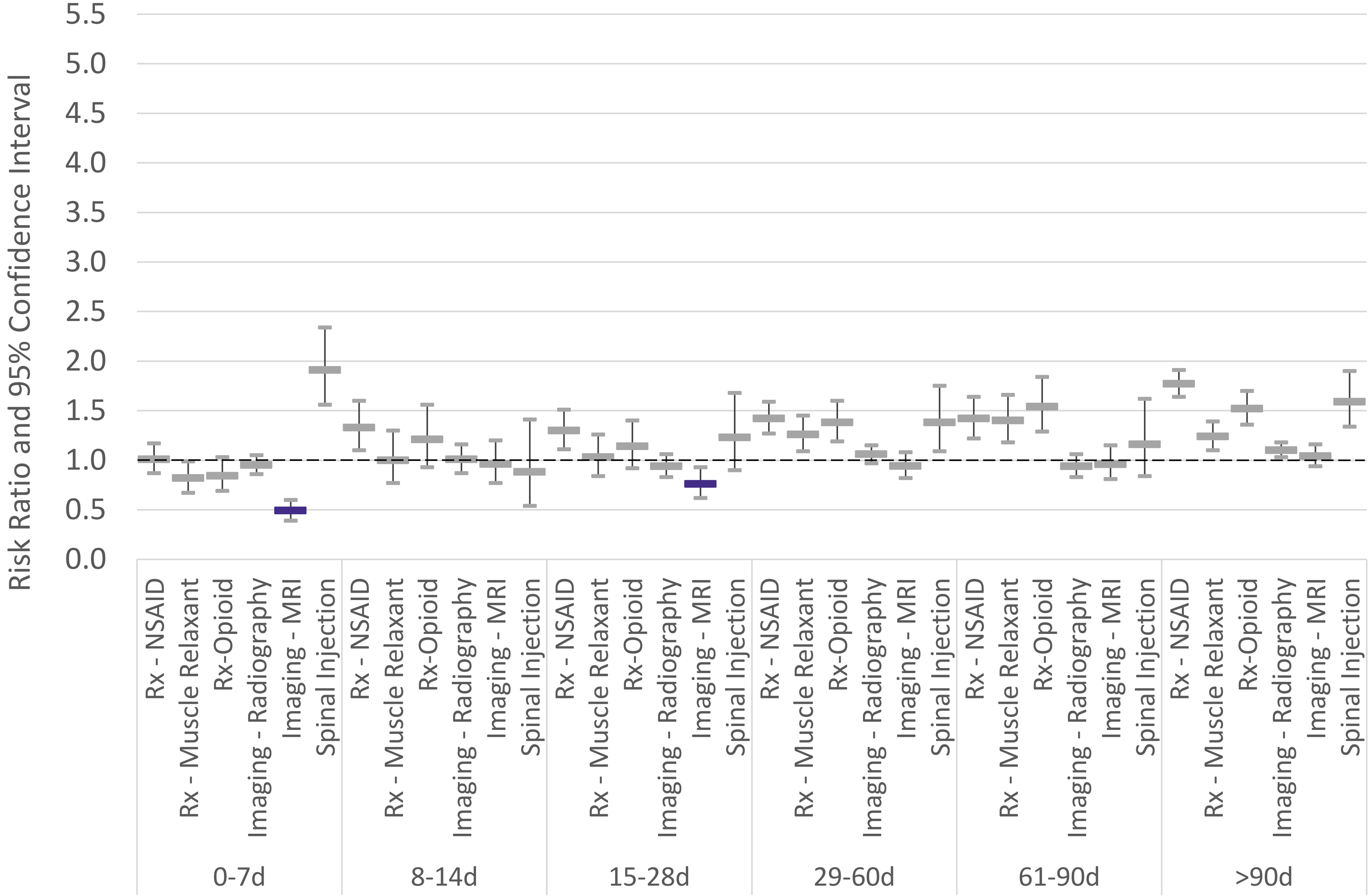


**Supplemental Figure 2.** Individuals with non-surgical neck pain initially contacting a physician specialist. Risk ratio and 95% confidence interval for exposure to health care services based on timing of introduction of **chiropractic manipulation** compared to episodes without chiropractic manipulation
