## Supplement - Neck Pain - Figure 3 for "Low back pain service utilization and costs: association with timing of first-line services for individuals initially contacting a physician specialist. A retrospective cohort study"

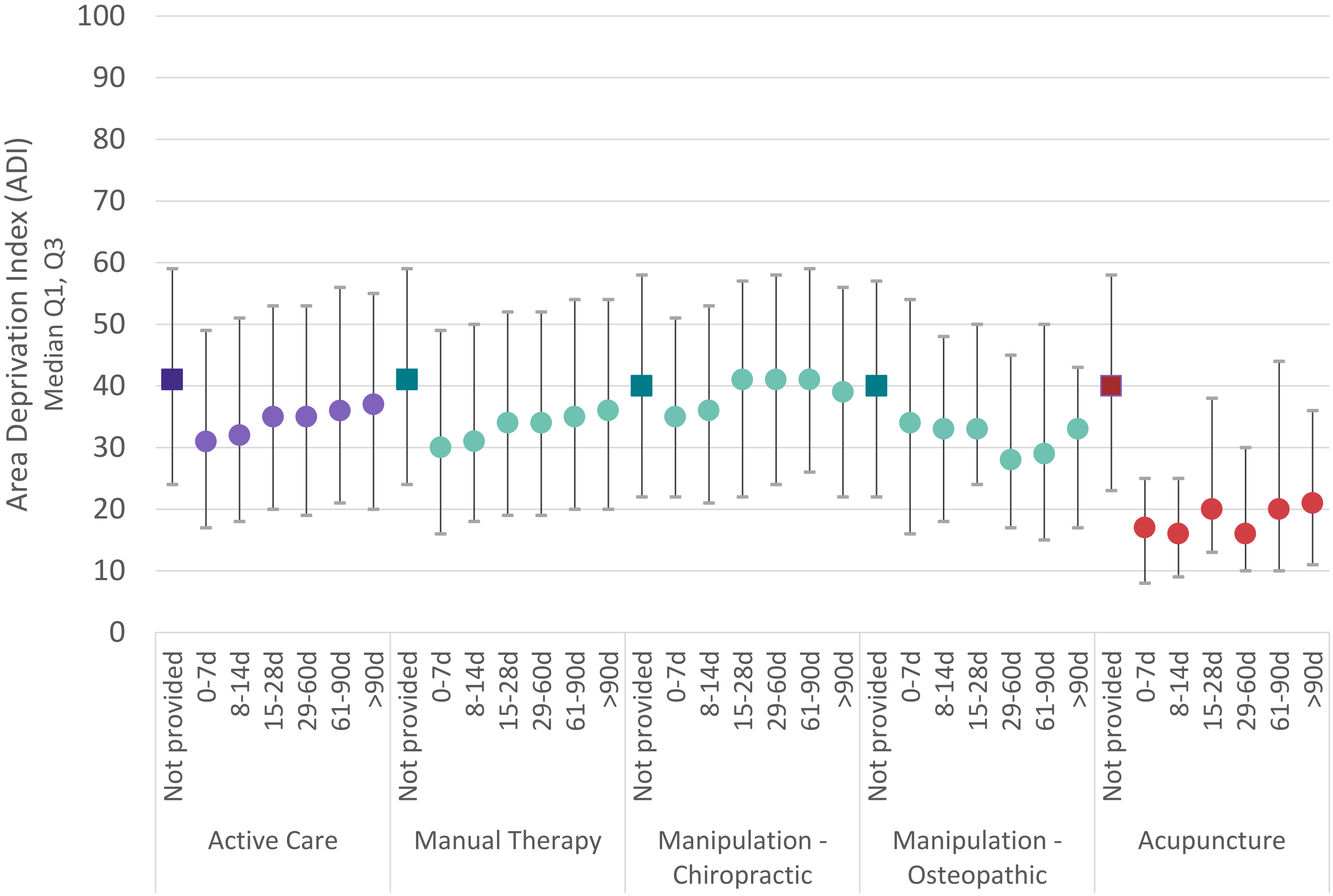


**Supplemental Figure 3.** For individuals with neck pain initially contacting a physician specialist, Area Deprivation Index (ADI) of the individual's home address zip code associated the number of days (d) into an episode when first line services are initially introduced
