## Supplement - Neck Pain - Figure 4 for "Low back pain service utilization and costs: association with timing of first-line services for individuals initially contacting a physician specialist. A retrospective cohort study"

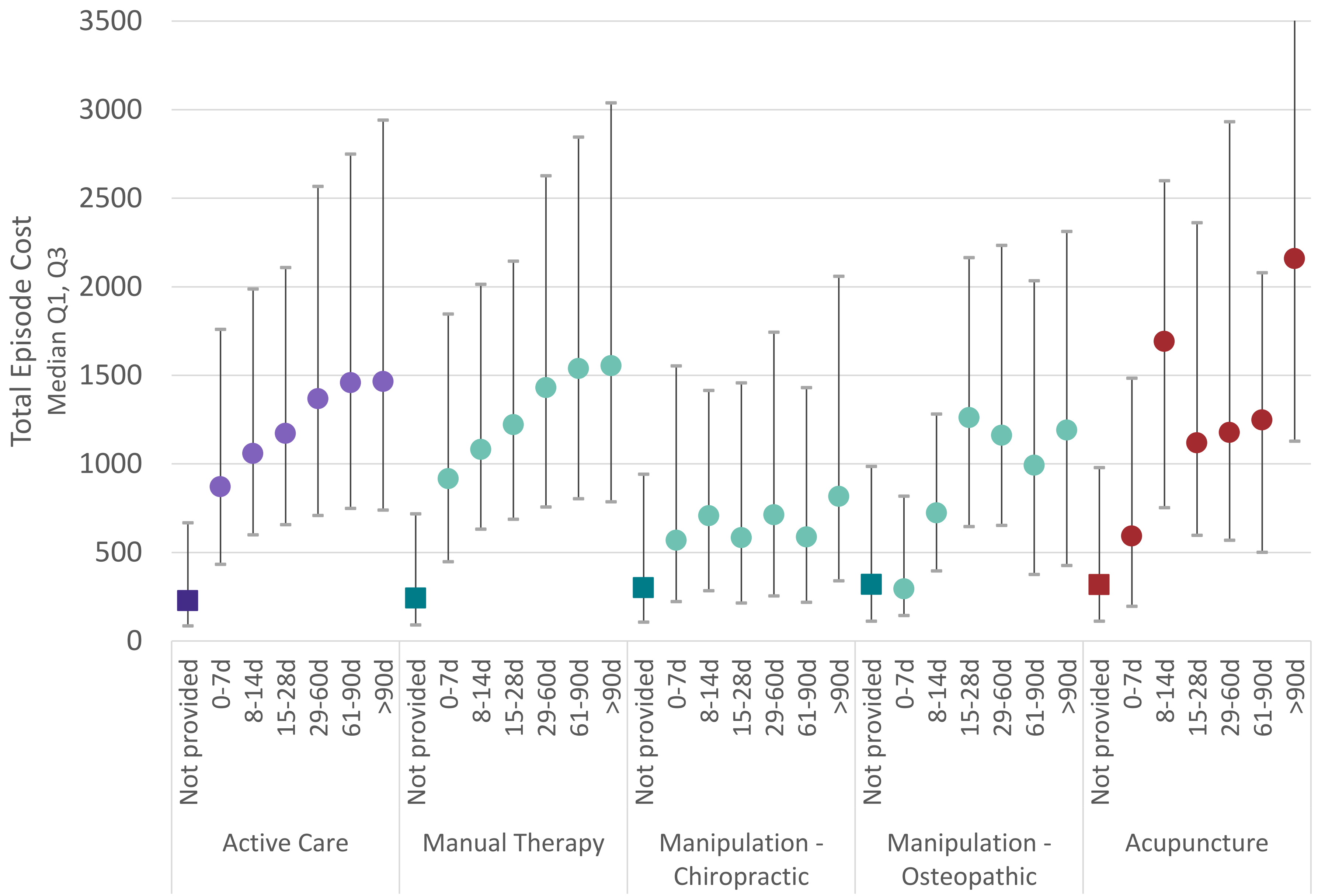


**Supplemental Figure 4.** For individuals with neck pain initially contacting a physician specialist, total episode cost associated with number of days (d) into an episode when first line services are initially introduced
