## Supplement - Neck Pain - Risk Ratio for "Low back pain service utilization and costs: association with timing of first-line services for individuals initially contacting a physician specialist. A retrospective cohort study"

| Neck Pain Supplement Risk Ratio - Individuals with non-surgical neck pain initially contacting a Specialist - risk ratio and 95% confidence interval for service exposure based on timing of introduction of first line services compared to if service not introduced | | | | | | | | | | | | | |
|---|---|---|---|---|---|---|---|---|---|---|---|---|---|
| | First Line | | | | | Passive Therapy | Second Line | | | | Third Line | | |
| | AC | MT | CMT | OMT | Acu | | Rx - NSAID | Rx - MM Relaxant | Imaging - Radiography | Imaging - MRI | Rx-Opioid | Spinal Injection | Imaging-CT |
| **Active Care (AC)** | | | | | | | | | | | | | |
| 0-7d | N/A | 45.47 (42.20, 49.00) | 3.06 (2.77, 3.38) | 2.69 (2.13, 3.40) | 6.81 (5.20, 8.93) | 16.89 (15.66, 18.20) | 1.14 (1.07, 1.22) | 0.83 (0.76, 0.91) | 1.18 (1.13, 1.23) | 0.90 (0.84, 0.97) | 0.60 (0.53, 0.67) | 1.17 (1.02, 1.33) | 0.43 (0.32, 0.57) |
| 8-14d | | 47.63 (44.12, 51.42) | 2.57 (2.22, 2.98) | 0.95 (0.56, 1.61) | 3.78 (2.41, 5.95) | 16.76 (15.35, 18.30) | 1.31 (1.21, 1.43) | 0.96 (0.86, 1.08) | 1.69 (1.62, 1.76) | 1.41 (1.30, 1.52) | 0.64 (0.55, 0.74) | 0.98 (0.81, 1.20) | 0.65 (0.47, 0.91) |
| 15-28d | | 47.54 (44.07, 51.29) | 2.79 (2.45, 3.19) | 0.75 (0.43, 1.31) | 2.63 (1.60, 4.32) | 17.62 (16.22, 19.15) | 1.17 (1.07, 1.27) | 1.04 (0.94, 1.15) | 1.49 (1.43, 1.56) | 1.87 (1.76, 1.99) | 0.73 (0.64, 0.83) | 1.26 (1.07, 1.48) | 0.69 (0.51, 0.92) |
| 29-60d | | 45.62 (42.25, 49.26) | 3.88 (3.47, 4.34) | 1.35 (0.89, 2.05) | 3.76 (2.45, 5.76) | 17.78 (16.37, 19.32) | 1.50 (1.40, 1.62) | 1.11 (1.01, 1.23) | 1.43 (1.37, 1.50) | 2.28 (2.16, 2.40) | 1.02 (0.91, 1.14) | 1.39 (1.19, 1.62) | 1.55 (1.27, 1.90) |
| 61-90d | | 42.84 (39.32, 46.67) | 6.10 (5.37, 6.93) | 1.97 (1.16, 3.34) | 6.00 (3.60, 9.98) | 18.04 (16.24, 20.05) | 1.59 (1.43, 1.77) | 1.34 (1.17, 1.54) | 1.34 (1.24, 1.44) | 2.27 (2.09, 2.46) | 1.36 (1.18, 1.57) | 2.02 (1.67, 2.45) | 1.70 (1.27, 2.28) |
| >90d | | 40.24 (37.13, 43.62) | 6.99 (6.40, 7.64) | 2.53 (1.81, 3.54) | 5.99 (4.11, 8.72) | 17.83 (16.36, 19.44) | 1.98 (1.85, 2.11) | 1.64 (1.51, 1.79) | 1.24 (1.18, 1.32) | 2.01 (1.88, 2.14) | 1.57 (1.43, 1.72) | 2.21 (1.94, 2.52) | 2.03 (1.67, 2.45) |
| **Manual Therapy (MT)** | | | | | | | | | | | | | |
| 0-7d | 15.66 (15.07, 16.27) | N/A | 2.13 (1.90, 2.40) | 1.42 (1.01, 1.98) | 9.15 (6.98, 12.01) | 12.41 (11.60, 13.27) | 1.12 (1.04, 1.21) | 0.85 (0.77, 0.94) | 1.17 (1.12, 1.23) | 0.89 (0.82, 0.97) | 0.62 (0.55, 0.70) | 1.19 (1.03, 1.38) | 0.40 (0.28, 0.56) |
| 8-14d | 16.07 (15.46, 16.71) | | 1.86 (1.57, 2.21) | 0.89 (0.50, 1.57) | 4.35 (2.71, 6.98) | 11.81 (10.87, 12.83) | 1.32 (1.20, 1.44) | 0.98 (0.86, 1.11) | 1.66 (1.58, 1.73) | 1.39 (1.28, 1.52) | 0.62 (0.52, 0.73) | 1.06 (0.86, 1.30) | 0.63 (0.44, 0.91) |
| 15-28d | 15.97 (15.37, 16.60) | | 1.88 (1.61, 2.19) | 0.60 (0.32, 1.12) | 4.10 (2.63, 6.38) | 12.42 (11.51, 13.39) | 1.15 (1.05, 1.25) | 0.99 (0.89, 1.11) | 1.48 (1.41, 1.55) | 1.81 (1.70, 1.94) | 0.71 (0.61, 0.82) | 1.24 (1.05, 1.48) | 0.59 (0.42, 0.83) |
| 29-60d | 15.81 (15.20, 16.44) | | 2.47 (2.16, 2.83) | 1.12 (0.70, 1.79) | 4.06 (2.58, 6.38) | 12.67 (11.75, 13.67) | 1.42 (1.31, 1.54) | 1.15 (1.04, 1.28) | 1.44 (1.37, 1.51) | 2.23 (2.11, 2.36) | 0.95 (0.84, 1.08) | 1.43 (1.21, 1.68) | 1.43 (1.14, 1.79) |
| 61-90d | 15.32 (14.65, 16.03) | | 3.93 (3.35, 4.62) | 1.24 (0.62, 2.48) | 7.68 (4.62, 12.76) | 12.29 (11.06, 13.66) | 1.55 (1.37, 1.74) | 1.40 (1.21, 1.62) | 1.35 (1.24, 1.46) | 2.24 (2.05, 2.45) | 1.36 (1.16, 1.59) | 2.11 (1.72, 2.60) | 1.85 (1.36, 2.51) |
| >90d | 14.54 (13.93, 15.18) | | 5.39 (4.90, 5.94) | 1.87 (1.25, 2.78) | 7.64 (5.28, 11.08) | 12.49 (11.52, 13.54) | 1.87 (1.74, 2.01) | 1.73 (1.58, 1.89) | 1.25 (1.17, 1.32) | 1.97 (1.83, 2.11) | 1.56 (1.41, 1.73) | 2.33 (2.03, 2.67) | 1.90 (1.53, 2.35) |
| **Manipulation - Chiropractic (CMT)** | | | | | | | | | | | | | |
| 0-7d | 3.06 (2.84, 3.30) | 2.25 (2.02, 2.52) | N/A | 1.08 (0.54, 2.16) | 4.01 (2.36, 6.83) | 6.27 (5.72, 6.88) | 1.01 (0.87, 1.17) | 0.82 (0.67, 0.99) | 0.95 (0.86, 1.05) | 0.49 (0.39, 0.60) | 0.84 (0.69, 1.03) | 1.91 (1.56, 2.34) | 0.55 (0.32, 0.94) |
| 8-14d | 2.81 (2.49, 3.18) | 2.37 (2.01, 2.79) | | 0.99 (0.32, 3.05) | 2.09 (0.67, 6.49) | 7.08 (6.26, 8.02) | 1.33 (1.10, 1.60) | 1.00 (0.77, 1.30) | 1.01 (0.87, 1.16) | 0.96 (0.77, 1.20) | 1.21 (0.93, 1.56) | 0.88 (0.54, 1.41) | 0.41 (0.16, 1.09) |
| 15-28d | 2.53 (2.28, 2.82) | 2.12 (1.84, 2.44) | | 1.03 (0.43, 2.46) | 3.48 (1.73, 6.99) | 6.70 (6.03, 7.44) | 1.30 (1.11, 1.51) | 1.03 (0.84, 1.26) | 0.94 (0.83, 1.06) | 0.76 (0.62, 0.93) | 1.14 (0.92, 1.40) | 1.23 (0.90, 1.68) | 0.96 (0.58, 1.58) |
| 29-60d | 2.69 (2.48, 2.92) | 2.38 (2.15, 2.65) | | 1.30 (0.70, 2.42) | 4.96 (3.09, 7.95) | 6.38 (5.84, 6.98) | 1.42 (1.27, 1.59) | 1.26 (1.09, 1.45) | 1.06 (0.97, 1.15) | 0.94 (0.82, 1.08) | 1.38 (1.19, 1.60) | 1.38 (1.09, 1.75) | 0.81 (0.53, 1.25) |
| 61-90d | 2.75 (2.48, 3.04) | 2.20 (1.91, 2.53) | | 0.42 (0.11, 1.69) | 5.39 (3.05, 9.54) | 6.31 (5.64, 7.06) | 1.42 (1.22, 1.64) | 1.40 (1.18, 1.66) | 0.94 (0.83, 1.06) | 0.96 (0.81, 1.15) | 1.54 (1.29, 1.84) | 1.16 (0.84, 1.62) | 0.86 (0.50, 1.47) |
| >90d | 2.78 (2.61, 2.96) | 2.46 (2.27, 2.67) | | 1.14 (0.67, 1.92) | 4.47 (3.00, 6.66) | 6.88 (6.42, 7.37) | 1.77 (1.64, 1.91) | 1.24 (1.10, 1.39) | 1.10 (1.03, 1.18) | 1.04 (0.94, 1.16) | 1.52 (1.36, 1.70) | 1.59 (1.34, 1.90) | 1.14 (0.85, 1.52) |
| **Manipulation - Osteopathic (OMT)** | | | | | | | | | | | | | |
| 0-7d | 1.22 (1.02, 1.46) | 0.79 (0.61, 1.02) | 0.60 (0.37, 0.97) | N/A | 2.99 (1.42, 6.28) | 1.32 (1.01, 1.71) | 0.57 (0.44, 0.74) | 0.67 (0.51, 0.88) | 0.29 (0.22, 0.38) | 0.26 (0.17, 0.38) | 0.42 (0.29, 0.62) | 1.70 (1.29, 2.24) | 0.14 (0.04, 0.58) |
| 8-14d | 1.65 (0.85, 3.20) | 1.35 (0.56, 3.22) | 0.79 (0.12, 5.31) | | N/A | 2.25 (0.94, 5.38) | 0.97 (0.41, 2.32) | 1.86 (0.96, 3.61) | 0.93 (0.52, 1.68) | 1.16 (0.55, 2.46) | N/A | 0.79 (0.12, 5.33) | N/A |
| 15-28d | 1.62 (0.94, 2.79) | 1.99 (1.15, 3.42) | 2.07 (0.83, 5.14) | | 23.46 (9.38, 58.65) | 2.21 (1.08, 4.51) | 0.64 (0.26, 1.58) | 1.42 (0.75, 2.72) | 0.79 (0.46, 1.35) | 0.91 (0.45, 1.86) | 1.16 (0.52, 2.57) | 3.63 (1.90, 6.92) | 0.99 (0.14, 6.81) |
| 29-60d | 2.56 (1.92, 3.41) | 1.83 (1.17, 2.85) | 2.14 (1.08, 4.25) | | 24.30 (12.13, 48.66) | 3.71 (2.52, 5.45) | 1.22 (0.77, 1.95) | 1.20 (0.69, 2.09) | 0.88 (0.60, 1.29) | 1.62 (1.12, 2.34) | 1.37 (0.79, 2.38) | 3.68 (2.25, 6.03) | 1.17 (0.30, 4.56) |
| 61-90d | 2.61 (1.91, 3.56) | 2.72 (1.90, 3.90) | 1.87 (0.82, 4.25) | | 8.50 (2.19, 32.97) | 3.21 (2.00, 5.15) | 1.04 (0.58, 1.84) | 2.07 (1.35, 3.15) | 0.76 (0.47, 1.22) | 0.11 (0.02, 0.76) | 0.84 (0.37, 1.91) | 4.13 (2.50, 6.84) | 1.44 (0.37, 5.55) |
| >90d | 2.26 (1.72, 2.97) | 1.53 (1.00, 2.34) | 2.01 (1.09, 3.70) | | 12.69 (5.42, 29.70) | 3.35 (2.35, 4.78) | 1.65 (1.20, 2.27) | 1.67 (1.14, 2.45) | 0.79 (0.56, 1.13) | 1.32 (0.91, 1.90) | 1.60 (1.04, 2.46) | 4.26 (2.91, 6.25) | 0.86 (0.22, 3.36) |
| **Acupuncture (Acu)** | | | | | | | | | | | | | |
| 0-7d | 2.60 (2.16, 3.14) | 3.04 (2.50, 3.71) | 1.48 (0.84, 2.60) | 10.06 (5.86, 17.29) | N/A | 6.44 (5.52, 7.52) | 0.57 (0.35, 0.93) | 0.42 (0.21, 0.81) | 0.38 (0.25, 0.59) | 0.31 (0.16, 0.61) | 0.18 (0.06, 0.55) | 1.45 (0.83, 2.55) | 0.26 (0.04, 1.80) |
| 8-14d | 3.41 (2.47, 4.71) | 5.19 (4.17, 6.47) | 4.56 (2.34, 8.92) | 9.48 (2.54, 35.38) | | 5.98 (4.02, 8.89) | 1.16 (0.54, 2.47) | 1.18 (0.49, 2.83) | 1.40 (0.94, 2.08) | 1.55 (0.85, 2.82) | 1.01 (0.36, 2.87) | 1.49 (0.40, 5.56) | N/A |
| 15-28d | 2.62 (1.87, 3.67) | 4.01 (3.07, 5.22) | 3.13 (1.62, 6.07) | 2.79 (0.40, 19.25) | | 4.47 (2.99, 6.69) | 0.82 (0.39, 1.69) | 0.35 (0.09, 1.33) | 0.75 (0.44, 1.26) | 1.31 (0.78, 2.20) | 0.40 (0.10, 1.52) | 0.88 (0.23, 3.37) | N/A |
| 29-60d | 3.00 (2.39, 3.77) | 3.48 (2.72, 4.44) | 4.89 (3.34, 7.16) | 5.08 (1.68, 15.31) | | 4.46 (3.26, 6.11) | 1.24 (0.80, 1.91) | 0.84 (0.44, 1.60) | 1.00 (0.72, 1.38) | 1.27 (0.84, 1.92) | 1.32 (0.78, 2.25) | 1.33 (0.58, 3.08) | 1.54 (0.51, 4.65) |
| 61-90d | 2.62 (1.86, 3.71) | 3.04 (2.10, 4.40) | 3.80 (2.09, 6.94) | 8.88 (3.02, 26.17) | | 3.40 (2.03, 5.68) | 1.16 (0.63, 2.11) | 0.55 (0.19, 1.62) | 0.48 (0.23, 0.98) | 1.11 (0.61, 2.02) | 1.47 (0.76, 2.84) | 0.93 (0.24, 3.57) | N/A |
| >90d | 3.09 (2.60, 3.68) | 3.68 (3.07, 4.41) | 5.24 (3.93, 6.98) | 4.21 (1.61, 11.02) | | 5.19 (4.18, 6.45) | 1.80 (1.39, 2.33) | 1.38 (0.95, 2.00) | 1.07 (0.84, 1.37) | 1.73 (1.33, 2.24) | 1.57 (1.08, 2.29) | 4.64 (3.41, 6.32) | 0.64 (0.16, 2.52) |

Confidence interval includes 1 for cells in red indicating risk is not different than the reference of if the specific service was not performed
