## Supplement - Neck Pain - State Summary for "Low back pain service utilization and costs: association with timing of first-line services for individuals initially contacting a physician specialist. A retrospective cohort study"

| Neck Pain Supplement - Episode count by home address state of individual with neck pain | | |
|---|---|---|
| State | Episodes | % |
| Total | 55025 | 100.00% |
| FL | 8056 | 14.64% |
| TX | 6864 | 12.47% |
| NY | 2954 | 5.37% |
| CA | 2576 | 4.68% |
| GA | 2382 | 4.33% |
| MD | 2379 | 4.32% |
| NC | 2158 | 3.92% |
| IL | 2028 | 3.69% |
| OH | 1964 | 3.57% |
| VA | 1932 | 3.51% |
| CO | 1788 | 3.25% |
| LA | 1699 | 3.09% |
| MO | 1594 | 2.90% |
| AZ | 1583 | 2.88% |
| WI | 1435 | 2.61% |
| NJ | 1358 | 2.47% |
| MN | 1124 | 2.04% |
| PA | 1057 | 1.92% |
| IN | 993 | 1.80% |
| TN | 962 | 1.75% |
| CT | 750 | 1.36% |
| OK | 541 | 0.98% |
| MS | 512 | 0.93% |
| NE | 492 | 0.89% |
| MI | 468 | 0.85% |
| AR | 468 | 0.85% |
| SC | 452 | 0.82% |
| MA | 437 | 0.79% |
| WA | 406 | 0.74% |
| UT | 385 | 0.70% |
| KY | 330 | 0.60% |
| IA | 300 | 0.55% |
| AL | 294 | 0.53% |
| RI | 266 | 0.48% |
| KS | 263 | 0.48% |
| NV | 262 | 0.48% |
| OR | 231 | 0.42% |
| DC | 223 | 0.41% |
| NM | 94 | 0.17% |
| WV | 92 | 0.17% |
| NH | 80 | 0.15% |
| VI | 58 | 0.11% |
| DE | 51 | 0.09% |
| ID | 51 | 0.09% |
| WY | 43 | 0.08% |
| ND | 37 | 0.07% |
| ME | 25 | 0.05% |
| MT | 18 | 0.03% |
| SD | 15 | 0.03% |
| PR | 9 | 0.02% |
| HI | 9 | 0.02% |
| AK | 6 | 0.01% |
| VT | 5 | 0.01% |
| Unknown | 466 | 0.85% |
