## Supplement - Neck Pain - Table 1 for "Low back pain service utilization and costs: association with timing of first-line services for individuals initially contacting a physician specialist. A retrospective cohort study"

| Neck Pain Supplement - Table 1 - Cohort characteristics - % or Median (Q1, Q3) | |
|---|---|
| # of Individuals | 50714 |
| # of Episodes | 55025 |
| # of Specialist health care providers (HCP) | 23356 |
| Total cost ($) | 53574827 |
| **Individuals with neck pain** | |
| % Female | 60.4% |
| Age | 49 (40, 56) |
| ERG® risk score | 2.4 (1.2, 4.3) |
| **Individual home address zip code population attributes** | |
| % non-Hispanic White (NHW) | 69.5% (49.6%, 82.7%) |
| Area Deprivation Index (ADI) | 40 (22, 57) |
| Household Adjusted Gross Income (AGI) ($) | 71852 (53384, 105947) |
| HCP per 1000 - DC | 0.23 (0.10, 0.43) |
| HCP per 1000 - PT | 0.19 (0.05, 0.44) |
| HCP per 1000 - LAc | 0.00 (0.00, 0.04) |
| **Episode attributes** | |
| Total cost ($) | 321 (113, 989) |
| # of HCP Seen | 2 (1, 3) |
| Episode duration - days (d) | 38 (1, 147) |
| Clean period - before initial episode (d) | 599 (382, 844) |
| Clean period - between sequential episodes (d) | 225 (126, 365) |
| Clean period - after final episode (d) | 447 (278, 721) |
