## Supplement - Neck Pain - Table 2 for "Low back pain service utilization and costs: association with timing of first-line services for individuals initially contacting a physician specialist. A retrospective cohort study"

Neck Pain Supplement - Table 2 - Non-surgical neck pain initially contacting specialist - episodic service use by number of days (d) into episode when first-line service first provided

| | Episodes | | First Line | | | | | Passive Therapy | Second Line | | | | Thirs Line | | |
|---|---|---|---|---|---|---|---|---|---|---|---|---|---|---|---|
| | Count | % | AC | MT | CMT | OMT | Acu | | Rx - NSAID | Rx - MM Relaxant | Imaging - Radiography | Imaging - MRI | Rx-Opioid | Spinal Injection | Imaging-CT |
| *Total* | 55025 | 100.0% | 19.3% | 15.7% | 6.7% | 1.1% | 0.6% | 9.5% | 21.7% | 16.9% | 39.2% | 22.5% | 14.8% | 6.8% | 3.5% |
| *Any First Line Service* | | | | | | | | | | | | | | | |
| **No First Line - reference** | **41639** | **75.7%** | **0.0%** | **0.0%** | **0.0%** | **0.0%** | **0.0%** | **0.3%** | **19.9%** | **16.6%** | **36.9%** | **20.4%** | **15.0%** | **6.1%** | **3.5%** |
| 0-7d | 4246 | 7.7% | 80.8% | 65.9% | 19.9% | 10.0% | 3.5% | 36.2% | 21.4% | 13.8% | 39.3% | 16.2% | 9.5% | 7.8% | 1.4% |
| 8-14d | 1747 | 3.2% | 88.6% | 73.7% | 18.4% | 1.5% | 1.6% | 38.8% | 26.1% | 16.4% | 59.5% | 28.2% | 10.5% | 5.7% | 2.3% |
| 15-28d | 2135 | 3.9% | 84.3% | 70.0% | 22.8% | 1.4% | 1.6% | 39.8% | 24.1% | 17.3% | 50.7% | 34.7% | 12.0% | 8.1% | 2.4% |
| 29-60d | 2208 | 4.0% | 79.2% | 64.3% | 30.5% | 1.9% | 2.4% | 39.0% | 29.4% | 19.2% | 49.5% | 40.6% | 17.1% | 8.8% | 4.9% |
| 61-90d | 983 | 1.8% | 72.0% | 56.6% | 41.1% | 2.5% | 3.0% | 38.1% | 30.8% | 23.7% | 43.4% | 37.7% | 22.3% | 11.5% | 5.0% |
| >90d | 2067 | 3.8% | 66.3% | 51.4% | 45.6% | 2.5% | 2.5% | 37.4% | 38.9% | 25.4% | 43.6% | 34.3% | 24.0% | 12.3% | 6.4% |
| *Active Care (AC)* | | | | | | | | | | | | | | | |
| **Not Provided - reference** | **44421** | **80.7%** | **0.0%** | **1.6%** | **4.3%** | **0.9%** | **0.4%** | **2.3%** | **20.2%** | **16.7%** | **36.6%** | **20.1%** | **15.2%** | **6.3%** | **3.4%** |
| 0-7d | 3177 | 5.8% | 100.0% | 74.7% | 13.2% | 2.5% | 2.4% | 38.4% | 23.0% | 13.8% | 43.2% | 18.2% | 9.1% | 7.3% | 1.5% |
| 8-14d | 1560 | 2.8% | 100.0% | 78.3% | 11.1% | 0.9% | 1.3% | 38.1% | 26.5% | 16.0% | 61.8% | 28.3% | 9.7% | 6.2% | 2.2% |
| 15-28d | 1820 | 3.3% | 100.0% | 78.1% | 12.0% | 0.7% | 0.9% | 40.1% | 23.5% | 17.4% | 54.7% | 37.6% | 11.0% | 7.9% | 2.4% |
| 29-60d | 1794 | 3.3% | 100.0% | 75.0% | 16.7% | 1.3% | 1.3% | 40.5% | 30.3% | 18.5% | 52.4% | 45.8% | 15.4% | 8.7% | 5.4% |
| 61-90d | 750 | 1.4% | 100.0% | 70.4% | 26.3% | 1.9% | 2.1% | 41.1% | 32.1% | 22.4% | 48.9% | 45.6% | 20.7% | 12.7% | 5.9% |
| > 90d | 1503 | 2.7% | 100.0% | 66.1% | 30.1% | 2.4% | 2.1% | 40.6% | 39.9% | 27.4% | 45.6% | 40.4% | 23.8% | 13.8% | 7.0% |
| *Manual Therapy (MT)* | | | | | | | | | | | | | | | |
| **Not Provided - reference** | **46411** | **84.3%** | **5.9%** | **0.0%** | **5.3%** | **1.1%** | **0.3%** | **3.4%** | **20.5%** | **16.6%** | **37.1%** | **20.6%** | **15.1%** | **6.3%** | **3.5%** |
| 0-7d | 2392 | 4.3% | 91.8% | 100.0% | 11.2% | 1.5% | 3.1% | 42.2% | 23.1% | 14.2% | 43.5% | 18.3% | 9.4% | 7.5% | 1.4% |
| 8-14d | 1275 | 2.3% | 94.2% | 100.0% | 9.8% | 0.9% | 1.5% | 40.2% | 27.1% | 16.2% | 61.5% | 28.6% | 9.3% | 6.7% | 2.2% |
| 15-28d | 1568 | 2.8% | 93.6% | 100.0% | 9.9% | 0.6% | 1.4% | 42.2% | 23.5% | 16.5% | 54.8% | 37.2% | 10.7% | 7.8% | 2.0% |
| 29-60d | 1511 | 2.7% | 92.7% | 100.0% | 13.0% | 1.2% | 1.4% | 43.1% | 29.3% | 19.2% | 53.3% | 45.9% | 14.4% | 9.0% | 5.0% |
| 61-90d | 608 | 1.1% | 89.8% | 100.0% | 20.7% | 1.3% | 2.6% | 41.8% | 31.7% | 23.4% | 50.0% | 46.1% | 20.6% | 13.3% | 6.4% |
| > 90d | 1260 | 2.3% | 85.2% | 100.0% | 28.4% | 2.0% | 2.6% | 42.5% | 38.3% | 28.8% | 46.2% | 40.4% | 23.7% | 14.7% | 6.6% |
| *Manipulation - Chiropractic (CMT)* | | | | | | | | | | | | | | | |
| **Not Provided - reference** | **51350** | **93.3%** | **17.2%** | **14.4%** | **0.0%** | **1.1%** | **0.5%** | **6.9%** | **21.1%** | **16.8%** | **39.2%** | **22.7%** | **14.6%** | **6.5%** | **3.5%** |
| 0-7d | 679 | 1.2% | 52.7% | 32.4% | 100.0% | 1.2% | 2.1% | 43.2% | 21.2% | 13.7% | 37.3% | 11.0% | 12.2% | 12.5% | 1.9% |
| 8-14d | 279 | 0.5% | 48.4% | 34.1% | 100.0% | 1.1% | 1.1% | 48.7% | 28.0% | 16.8% | 39.4% | 21.9% | 17.6% | 5.7% | 1.4% |
| 15-28d | 447 | 0.8% | 43.6% | 30.4% | 100.0% | 1.1% | 1.8% | 46.1% | 27.3% | 17.2% | 36.9% | 17.2% | 16.6% | 8.1% | 3.4% |
| 29-60d | 706 | 1.3% | 46.3% | 34.3% | 100.0% | 1.4% | 2.5% | 43.9% | 29.9% | 21.1% | 41.4% | 21.4% | 20.1% | 9.1% | 2.8% |
| 61-90d | 433 | 0.8% | 47.3% | 31.6% | 100.0% | 0.5% | 2.8% | 43.4% | 29.8% | 23.6% | 36.7% | 21.9% | 22.4% | 7.6% | 3.0% |
| > 90d | 1131 | 2.1% | 47.8% | 35.4% | 100.0% | 1.2% | 2.3% | 47.3% | 37.3% | 20.8% | 43.1% | 23.7% | 22.2% | 10.4% | 4.0% |
| *Manipulation - Osteopathic (OMT)* | | | | | | | | | | | | | | | |
| **Not Provided - reference** | **54423** | **98.9%** | **19.2%** | **15.6%** | **6.7%** | **0.0%** | **0.6%** | **9.4%** | **21.7%** | **16.9%** | **39.5%** | **22.7%** | **14.9%** | **6.7%** | **3.5%** |
| 0-7d | 398 | 0.7% | 23.4% | 12.3% | 4.0% | 100.0% | 1.8% | 12.3% | 12.3% | 11.3% | 11.6% | 5.8% | 6.3% | 11.3% | 0.5% |
| 8-14d | 19 | 0.0% | 31.6% | 21.1% | 5.3% | 100.0% | 0.0% | 21.1% | 21.1% | 31.6% | 36.8% | 26.3% | 0.0% | 5.3% | 0.0% |
| 15-28d | 29 | 0.1% | 31.0% | 31.0% | 13.8% | 100.0% | 13.8% | 20.7% | 13.8% | 24.1% | 31.0% | 20.7% | 17.2% | 24.1% | 3.4% |
| 29-60d | 49 | 0.1% | 49.0% | 28.6% | 14.3% | 100.0% | 14.3% | 34.7% | 26.5% | 20.4% | 34.7% | 36.7% | 20.4% | 24.5% | 4.1% |
| 61-90d | 40 | 0.1% | 50.0% | 42.5% | 12.5% | 100.0% | 5.0% | 30.0% | 22.5% | 35.0% | 30.0% | 2.5% | 12.5% | 27.5% | 5.0% |
| > 90d | 67 | 0.1% | 43.3% | 23.9% | 13.4% | 100.0% | 7.5% | 31.3% | 35.8% | 28.4% | 31.3% | 29.9% | 23.9% | 28.4% | 3.0% |
| *Acupuncture (Acu)* | | | | | | | | | | | | | | | |
| **Not Provided - reference** | **54680** | **99.4%** | **19.1%** | **15.4%** | **6.6%** | **1.1%** | **0.0%** | **9.2%** | **21.6%** | **17.0%** | **39.3%** | **22.5%** | **14.9%** | **6.7%** | **3.5%** |
| 0-7d | 113 | 0.2% | 49.6% | 46.9% | 9.7% | 10.6% | 100.0% | 59.3% | 12.4% | 7.1% | 15.0% | 7.1% | 2.7% | 9.7% | 0.9% |
| 8-14d | 20 | 0.0% | 65.0% | 80.0% | 30.0% | 10.0% | 100.0% | 55.0% | 25.0% | 20.0% | 55.0% | 35.0% | 15.0% | 10.0% | 0.0% |
| 15-28d | 34 | 0.1% | 50.0% | 61.8% | 20.6% | 2.9% | 100.0% | 41.2% | 17.6% | 5.9% | 29.4% | 29.4% | 5.9% | 5.9% | 0.0% |
| 29-60d | 56 | 0.1% | 57.1% | 53.6% | 32.1% | 5.4% | 100.0% | 41.1% | 26.8% | 14.3% | 39.3% | 28.6% | 19.6% | 8.9% | 5.4% |
| 61-90d | 32 | 0.1% | 50.0% | 46.9% | 25.0% | 9.4% | 100.0% | 31.2% | 25.0% | 9.4% | 18.8% | 25.0% | 21.9% | 6.2% | 0.0% |
| > 90d | 90 | 0.2% | 58.9% | 56.7% | 34.4% | 4.4% | 100.0% | 47.8% | 38.9% | 23.3% | 42.2% | 38.9% | 23.3% | 31.1% | 2.2% |

Cells with red text denote that service usage was not significantly different from the referent of no first line service (Fishers's Exact p=>0.001)
Cells with red text denote that service usage was significantly different from the referent of no first line service (Fishers's Exact p=>0.001)

AC=Active Care, MT=Manual Therapy, CMT=Chiropractic Manipulative Treatment, OMT=Osteopathic Manipulative Treatment, Acu=Acupuncture
