## Supplement - Neck Pain - Table 3 for "Low back pain service utilization and costs: association with timing of first-line services for individuals initially contacting a physician specialist. A retrospective cohort study"

**Neck Pain Supplement - Table 3 - Individual, local population and episode attributes associated with individuals with neck pain initially contacting a specialist by timing of incorporation of first line services**

| Timing Days (d) | Specialists | | Episodes | | Episode Attributes Median (Q1, Q3) | | Individual Attributes Median (Q1, Q3) | | Individual Home Address Zip Code Population Attributes Median (Q1, Q3) | | | | | |
|---|---|---|---|---|---|---|---|---|---|---|---|---|---|---|
| | | | | | | | | | % NHW | ADI | AGI | HCP per 1000 population | | |
| | Count | % | Count | % | Total Cost | Duration | Age | ERG® Risk | | | | DC | PT | LAc |
| ***Total*** | 23356 | 100.0% | 55025 | 100.0% | 321 (113, 989) | 38 (1, 147) | 49 (40, 56) | 2.4 (1.2, 4.3) | 69.5% (49.6%, 82.7%) | 40 (22, 57) | 71852 (53384, 105947) | 0.23 (0.10, 0.43) | 0.19 (0.05, 0.44) | 0.00 (0.00, 0.04) |
| ***Any First Line Service*** | | | | | | | | | | | | | | |
| **Not provided** | 14967 | 64.1% | 41639 | 75.7% | 218 (80, 641) | 18 (1, 118) | 49 (40, 57) | 2.4 (1.2, 4.3) | 68.9% (49.0%, 82.5%) | 41 (24, 59) | 70190 (52346, 103105) | 0.22 (0.09, 0.42) | 0.18 (0.05, 0.42) | 0.00 (0.00, 0.03) |
| **0-7d** | 2728 | 11.7% | 4246 | 7.7% | 752 (335, 1596) | 51 (17, 149) | 48 (38, 55) | 1.9 (0.9, 3.5) | 71.2% (51.4%, 83.1%) | 32 (18, 50) | 80331 (58722, 121058) | 0.26 (0.12, 0.47) | 0.23 (0.08, 0.50) | 0.00 (0.00, 0.06) |
| **8-14d** | 1488 | 6.4% | 1747 | 3.2% | 995 (552, 1906) | 56 (31, 124) | 49 (39, 56) | 1.9 (1.0, 3.5) | 69.5% (51.6%, 82.6%) | 33 (18, 51) | 80798 (59121, 118664) | 0.24 (0.11, 0.47) | 0.21 (0.06, 0.48) | 0.00 (0.00, 0.05) |
| **15-28d** | 1891 | 8.1% | 2135 | 3.9% | 1092 (542, 2007) | 68 (39, 148) | 49 (39, 56) | 2.2 (1.1, 3.8) | 71.2% (50.7%, 83.5%) | 36 (21, 54) | 76030 (56202, 111676) | 0.26 (0.11, 0.46) | 0.21 (0.06, 0.48) | 0.00 (0.00, 0.05) |
| **29-60d** | 2001 | 8.6% | 2208 | 4.0% | 1201 (542, 2310) | 96 (60, 188) | 49 (40, 57) | 2.5 (1.3, 4.1) | 70.8% (52.9%, 83.6%) | 37 (20, 54) | 75626 (55724, 109534) | 0.26 (0.12, 0.46) | 0.19 (0.06, 0.46) | 0.00 (0.00, 0.04) |
| **61-90d** | 946 | 4.1% | 983 | 1.8% | 1073 (430, 2268) | 136 (92, 229) | 49 (41, 56) | 2.7 (1.4, 4.9) | 72.0%) (56.4%, 84.4%) | 39 (22, 58) | 74089 (54159, 107297) | 0.25 (0.12, 0.44) | 0.20 (0.06, 0.45) | 0.00 (0.00, 0.04) |
| **>90d** | 1941 | 8.3% | 2067 | 3.8% | 1126 (490, 2471) | 239 (175, 319) | 50 (41, 57) | 3.0 (1.7, 4.8) | 72.0% (52.7%, 84.4%) | 39 (22, 56) | 74529 (55070, 107509) | 0.24 (0.11, 0.46) | 0.19 (0.05, 0.45) | 0.00 (0.00, 0.04) |
| ***Active Care*** | | | | | | | | | | | | | | |
| **Not provided** | 16626 | 71.2% | 44421 | 80.7% | 227 (85, 667) | 23 (1, 128) | 49 (40, 56) | 2.4 (1.2, 4.3) | 69.5% (49.6%, 82.9%) | 41 (24, 59) | 70326 (52509, 103138) | 0.22 (0.10, 0.42) | 0.18 (0.05, 0.43) | 0.00 (0.00, 0.03) |
| **0-7d** | 2148 | 9.2% | 3177 | 5.8% | 870 (433, 1759) | 53 (22, 148) | 48 (38, 56) | 1.9 (0.9, 3.5) | 69.5% (49.8%, 81.9%) | 31 (17, 49) | 82317 (59468, 125544) | 0.26 (0.12, 0.48) | 0.23 (0.08, 0.49) | 0.00 (0.00, 0.06) |
| **8-14d** | 1317 | 5.6% | 1560 | 2.8% | 1058 (600, 1987) | 54 (32, 120) | 48 (39, 56) | 1.9 (0.9, 3.5) | 68.6% (50.0%, 82.2%) | 32 (18, 51) | 81427 (59193, 121336) | 0.23 (0.11, 0.46) | 0.20 (0.06, 0.49) | 0.00 (0.00, 0.05) |
| **15-28d** | 1602 | 6.9% | 1820 | 3.3% | 1171 (657, 2108) | 65 (40, 136) | 49 (39, 56) | 2.1 (1.1, 3.7) | 69.8% (49.6%, 82.5%) | 35 (20, 53) | 77890 (57233, 114745) | 0.26 (0.11, 0.46) | 0.22 (0.08, 0.47) | 0.00 (0.00, 0.05) |
| **29-60d** | 1645 | 7.0% | 1794 | 3.3% | 1368 (709, 2567) | 93 (61, 169) | 50 (40, 57) | 2.5 (1.3, 4.2) | 69.5% (50.9%, 82.7%) | 35 (19, 53) | 77860 (57025, 113899) | 0.25 (0.12, 0.46) | 0.20 (0.06, 0.46) | 0.00 (0.00, 0.05) |
| **61-90d** | 725 | 3.1% | 750 | 1.4% | 1458 (749, 2749) | 133 (94, 222) | 50 (41, 57) | 2.8 (1.5, 4.9) | 70.8% (54.3%, 83.2%) | 36 (21, 56) | 78244 (54775, 115628) | 0.25 (0.11, 0.44) | 0.21 (0.06, 0.46) | 0.00 (0.00, 0.05) |
| **>90d** | 1433 | 6.1% | 1503 | 2.7% | 1465 (739, 2941) | 239 (176, 319) | 51 (41, 57) | 3.2 (1.8, 5.0) | 69.7% (48.9%, 82.6%) | 37 (20, 55) | 76441 (55491, 111183) | 0.24 (0.11, 0.46) | 0.19 (0.05, 0.45) | 0.00 (0.00, 0.04) |
| ***Manual Therapy*** | | | | | | | | | | | | | | |
| **Not provided** | 17639 | 75.5% | 46411 | 84.3% | 242 (91, 718) | 26 (1, 132) | 49 (40, 56) | 2.4 (1.2, 4.3) | 69.5% (49.6%, 82.8%) | 41 (24, 59) | 70538 (52619, 103559) | 0.22 (0.10, 0.42) | 0.18 (0.05, 0.43) | 0.00 (0.00, 0.03) |
| **0-7d** | 1686 | 7.2% | 2392 | 4.3% | 917 (448, 1846) | 55 (23, 155) | 48 (38, 55) | 1.9 (0.9, 3.5) | 68.4% (49.5%, 82.1%) | 30 (16, 49) | 83602 (59503, 128914) | 0.25 (0.12, 0.46) | 0.23 (0.08, 0.50) | 0.00 (0.00, 0.06) |
| **8-14d** | 1113 | 4.8% | 1275 | 2.3% | 1081 (632, 2014) | 54 (31, 120) | 49 (39, 56) | 1.9 (0.9, 3.4) | 69.7% (50.8%, 82.5%) | 31 (18, 50) | 81170 (59931, 121601) | 0.23 (0.11, 0.44) | 0.22 (0.08, 0.49) | 0.00 (0.00, 0.05) |
| **15-28d** | 1399 | 6.0% | 1568 | 2.8% | 1222 (687, 2145) | 67 (42, 141) | 49 (40, 56) | 2.1 (1.1, 3.8) | 69.6% (49.9%, 82.3%) | 34 (19, 52) | 78895 (57353, 117687) | 0.26 (0.11, 0.46) | 0.22 (0.06, 0.49) | 0.00 (0.00, 0.06) |
| **29-60d** | 1379 | 5.9% | 1511 | 2.7% | 1430 (756, 2627) | 92 (61, 170) | 50 (40, 57) | 2.4 (1.3, 4.1) | 69.6% (51.7%, 82.5%) | 34 (19, 52) | 80645 (58407, 116199) | 0.25 (0.13, 0.47) | 0.20 (0.07, 0.46) | 0.00 (0.00, 0.04) |
| **61-90d** | 590 | 2.5% | 608 | 1.1% | 1539 (803, 2845) | 130 (96, 222) | 50 (42, 57) | 2.9 (1.5, 4.6) | 71.0% (54.6%, 83.2%) | 35 (20, 54) | 78244 (55434, 115554) | 0.26 (0.13, 0.47) | 0.23 (0.07, 0.49) | 0.00 (0.00, 0.05) |
| **>90d** | 1211 | 5.2% | 1260 | 2.3% | 1555 (786, 3038) | 241 (177, 330) | 50 (41, 57) | 3.1 (1.7, 4.9) | 70.4% (48.7%, 82.9%) | 36 (20, 54) | 75599 (55831, 111676) | 0.23 (0.11, 0.45) | 0.19 (0.06, 0.46) | 0.00 (0.00, 0.04) |
| ***Manipulation - Chiropractic*** | | | | | | | | | | | | | | |
| **Not provided** | 20181 | 86.4% | 51350 | 93.3% | 300 (107, 942) | 33 (1, 132) | 49 (40, 57) | 2.4 (1.2, 4.3) | 69.0% (49.2%, 82.4%) | 40 (22, 58) | 71641 (53229, 105870) | 0.23 (0.10, 0.42) | 0.18 (0.05, 0.44) | 0.00 (0.00, 0.04) |
| **0-7d** | 513 | 2.2% | 679 | 1.2% | 569 (222, 1553) | 70 (17, 198) | 46 (36, 55) | 2.0 (0.9, 3.7) | 74.8% (57.3%, 86.5%) | 35 (22, 51) | 75167 (59628, 108161) | 0.28 (0.15, 0.50) | 0.24 (0.09, 0.54) | 0.00 (0.00, 0.05) |
| **8-14d** | 270 | 1.2% | 279 | 0.5% | 707 (284, 1414) | 80 (28, 225) | 48 (39, 55) | 2.3 (1.2, 4.0) | 70.8% (56.2%, 84.9%) | 36 (21, 53) | 76314 (56850, 111975) | 0.28 (0.14, 0.51) | 0.21 (0.06, 0.48) | 0.00 (0.00, 0.05) |
| **15-28d** | 444 | 1.9% | 447 | 0.8% | 583 (214, 1457) | 88 (36, 215) | 48 (38, 55) | 2.5 (1.3, 4.1) | 76.8% (59.1%, 87.4%) | 41 (22, 57) | 74276 (54746, 106061) | 0.28 (0.12, 0.50) | 0.20 (0.05, 0.47) | 0.00 (0.00, 0.03) |
| **29-60d** | 688 | 2.9% | 706 | 1.3% | 713 (255, 1744) | 122 (62, 238) | 47 (38, 54) | 2.2 (1.2, 4.0) | 74.6% (57.1%, 86.0%) | 41 (24, 58) | 70252 (53394, 100997) | 0.27 (0.12, 0.49) | 0.22 (0.07, 0.47) | 0.00 (0.00, 0.03) |
| **61-90d** | 430 | 1.8% | 433 | 0.8% | 588 (218, 1431) | 147 (91, 252) | 48 (38, 55) | 2.4 (1.3, 4.8) | 74.9% (60.1%, 86.6%) | 41 (26, 59) | 70216 (53509, 98219) | 0.25 (0.12, 0.45) | 0.19 (0.06, 0.38) | 0.00 (0.00, 0.03) |
| **>90d** | 1090 | 4.7% | 1131 | 2.1% | 816 (340, 2059) | 245 (179, 329) | 48 (39, 55) | 2.6 (1.5, 4.2) | 74.4% (56.9%, 86.3%) | 39 (22, 56) | 75074 (55045, 110329) | 0.26 (0.13, 0.48) | 0.20 (0.06, 0.47) | 0.00 (0.00, 0.04) |
| ***Manipulation - Osteopathic*** | | | | | | | | | | | | | | |
| **Not provided** | 23053 | 98.7% | 54423 | 98.9% | 319 (112, 986) | 38 (1, 146) | 49 (40, 56) | 2.4 (1.2, 4.3) | 69.4% (49.6%, 82.7%) | 40 (22, 57) | 71703 (53366, 105840) | 0.23 (0.10, 0.43) | 0.19 (0.05, 0.44) | 0.00 (0.00, 0.04) |
| **0-7d** | 169 | 0.7% | 398 | 0.7% | 295 (144, 818) | 32 (1, 120) | 47 (34, 56) | 1.7 (0.8, 3.4) | 74.5% (58.7%, 84.6%) | 34 (16, 54) | 80508 (55733, 117580) | 0.27 (0.13, 0.47) | 0.24 (0.08, 0.56) | 0.00 (0.00, 0.07) |
| **8-14d** | 17 | 0.1% | 19 | 0.0% | 724 (396, 1282) | 47 (18, 127) | 51 (45, 56) | 2.8 (1.1, 3.5) | 70.7% (59.1%, 77.5%) | 33 (18, 48) | 97673 (65177, 128992) | 0.38 (0.17, 0.49) | 0.32 (0.08, 0.90) | 0.03 (0.00, 0.08) |
| **15-28d** | 28 | 0.1% | 29 | 0.1% | 1261 (646, 2164) | 132 (42, 305) | 49 (44, 58) | 3.7 (1.8, 4.7) | 80.9% (68.5%, 85.2%) | 33 (24, 50) | 84642 (58341, 108407) | 0.23 (0.16, 0.42) | 0.13 (0.05, 0.39) | 0.00 (0.00, 0.05) |
| **29-60d** | 45 | 0.2% | 49 | 0.1% | 1162 (653, 2234) | 121 (55, 229) | 47 (39, 56) | 2.6 (1.5, 4.3) | 67.2% (54.5%, 80.5%) | 28 (17, 45) | 88958 (61889, 111705) | 0.36 (0.16, 0.46) | 0.32 (0.12, 0.57) | 0.03 (0.00, 0.09) |
| **61-90d** | 38 | 0.2% | 40 | 0.1% | 993 (376, 2034) | 150 (101, 223) | 47 (36, 57) | 2.7 (1.3, 4.1) | 73.0% (53.0%, 82.0%) | 29 (15, 50) | 77336 (58153, 120184) | 0.25 (0.15, 0.40) | 0.19 (0.06, 0.42) | 0.00 (0.00, 0.07) |
| **>90d** | 63 | 0.3% | 67 | 0.1% | 1191 (426, 2313) | 273 (233, 336) | 49 (38, 57) | 2.7 (1.8, 6.0) | 72.6% (57.2%, 81.2%) | 33 (17, 43) | 88143 (70171, 126760) | 0.26 (0.16, 0.52) | 0.18 (0.09, 0.64) | 0.01 (0.00, 0.11) |
| ***Acupuncture*** | | | | | | | | | | | | | | |
| **Not provided** | 23098 | 98.9% | 54680 | 99.4% | 318 (112, 979) | 38 (1, 146) | 49 (40, 56) | 2.4 (1.2, 4.3) | 69.5% (49.7%, 82.8%) | 40 (23, 58) | 71651 (53340, 105740) | 0.23 (0.10, 0.43) | 0.19 (0.05, 0.44) | 0.00 (0.00, 0.04) |
| **0-7d** | 52 | 0.2% | 113 | 0.2% | 592 (196, 1484) | 43 (4, 118) | 46 (38, 54) | 1.5 (0.7, 3.0) | 58.7% (42.2%, 72.5%) | 17 (8, 25) | 100734 (62720, 188389) | 0.27 (0.11, 0.45) | 0.37 (0.16, 0.77) | 0.05 (0.00, 0.14) |
| **8-14d** | 20 | 0.1% | 20 | 0.0% | 1691 (752, 2599) | 104 (23, 170) | 46 (41, 50) | 1.6 (1.0, 2.1) | 57.1% (49.6%, 70.7%) | 16 (9, 25) | 128418 (98444, 159903) | 0.29 (0.09, 0.52) | 0.43 (0.20, 0.49) | 0.05 (0.02, 0.19) |
| **15-28d** | 33 | 0.1% | 34 | 0.1% | 1118 (597, 2362) | 73 (52, 230) | 45 (36, 55) | 2.6 (1.4, 3.3) | 57.6% (43.1%, 78.3%) | 20 (13, 38) | 97394 (70989, 139967) | 0.29 (0.13, 0.47) | 0.30 (0.09, 0.94) | 0.04 (0.00, 0.19) |
| **29-60d** | 52 | 0.2% | 56 | 0.1% | 1177 (569, 2932) | 103 (62, 242) | 46 (39, 53) | 1.8 (1.0, 2.9) | 65.7% (45.8%, 76.9%) | 16 (10, 30) | 99817 (66631, 184460) | 0.29 (0.12, 0.51) | 0.26 (0.15, 0.78) | 0.04 (0.00, 0.16) |
| **61-90d** | 32 | 0.1% | 32 | 0.1% | 1247 (501, 2079) | 117 (97, 192) | 47 (38, 56) | 2.4 (1.6, 4.0) | 67.1% (55.0%, 79.3%) | 20 (10, 44) | 90813 (66884, 207134) | 0.21 (0.12, 0.35) | 0.29 (0.06, 0.59) | 0.00 (0.00, 0.06) |
| **>90d** | 89 | 0.4% | 90 | 0.2% | 2158 (1128, 4399) | 251 (170, 360) | 46 (38, 55) | 2.7 (1.3, 4.4) | 60.5% (45.0%, 77.6%) | 21 (11, 36) | 95408 (73135, 145533) | 0.27 (0.14, 0.50) | 0.21 (0.07, 0.57) | 0.04 (0.00, 0.12) |

ERG®=Episode Risk Group, NHW=Non-Hispanic White, ADI=Area Deprivation Index, AGI=Adjusted Gross Income, HCP=Health Care Provider, DC=Doctor of Chiropractic, PT=Physical Therapist, LAc=Licensed Acupuncturist

Cells with red text denote that the effect of first line service timing on measured attributes was found not to be significantly different from that of Not Provided reference - (Mann-Whitney U $p > 0.001$)

Cells with black text denote that the effect of first line service timing on measured attributes was found to be significantly different from that of Not Provided reference (ref) - (Mann-Whitney U $p < 0.001$)
