## Supplement - Risk Ratio for "Low back pain service utilization and costs: association with timing of first-line services for individuals initially contacting a physician specialist. A retrospective cohort study"

| Supplement Risk Ratio - Individuals with non-surgical low back pain initially contacting a specialist - risk ratio and 95% confidence interval for service exposure based on timing of introduction of first line services compared to if service not introduced | | | | | | | | | | | | | |
|---|---|---|---|---|---|---|---|---|---|---|---|---|---|
| | First Line | | | | | Passive Therapy | Second Line | | | | Third Line | | |
| | Active Care | Manual Therapy | Manipulation | | Acupuncture | | Rx | | Imaging | | Rx-Opioid | Spinal Injection | Imaging-CT |
| | | | Chiropractic | Osteopathic | | | NSAID | MM Relaxant | Radiography | MRI | | | |
| **Any First Line** | | | | | | | | | | | | | |
| 0-7d | | | | | | 151.22 (128.32, 178.22) | 1.01 (0.97, 1.05) | 0.88 (0.83, 0.93) | 1.17 (1.14, 1.20) | 0.96 (0.91, 1.00) | 0.59 (0.55, 0.63) | 0.84 (0.79, 0.90) | 0.71 (0.58, 0.85) |
| 8-14d | | | | | | 144.25 (121.77, 170.87) | 1.23 (1.17, 1.30) | 0.99 (0.91, 1.07) | 1.59 (1.55, 1.64) | 1.38 (1.30, 1.46) | 0.64 (0.59, 0.71) | 0.86 (0.78, 0.95) | 0.53 (0.39, 0.73) |
| 15-28d | | | | | | 141.57 (119.57, 167.62) | 1.18 (1.12, 1.25) | 1.07 (0.99, 1.15) | 1.47 (1.42, 1.51) | 1.82 (1.74, 1.91) | 0.83 (0.77, 0.90) | 1.02 (0.94, 1.11) | 0.71 (0.54, 0.92) |
| 29-60d | | | | | | 150.30 (127.10, 177.74) | 1.31 (1.24, 1.37) | 1.08 (1.00, 1.16) | 1.31 (1.27, 1.35) | 2.04 (1.96, 2.13) | 1.00 (0.93, 1.07) | 1.33 (1.24, 1.43) | 1.31 (1.09, 1.59) |
| 61-90d | | | | | | 153.13 (128.58, 182.37) | 1.47 (1.38, 1.57) | 1.20 (1.09, 1.32) | 1.29 (1.23, 1.35) | 1.85 (1.74, 1.97) | 1.23 (1.13, 1.33) | 1.51 (1.38, 1.66) | 1.58 (1.24, 2.02) |
| >90d | | | | | | 174.65 (147.96, 206.15) | 1.66 (1.60, 1.73) | 1.47 (1.39, 1.56) | 1.20 (1.16, 1.24) | 1.79 (1.72, 1.87) | 1.41 (1.34, 1.48) | 1.81 (1.71, 1.92) | 2.14 (1.85, 2.48) |
| **Active Care (AC)** | | | | | | | | | | | | | |
| 0-7d | | 52.53 (49.29, 55.98) | 2.53 (2.34, 2.75) | 3.09 (2.54, 3.76) | 4.06 (3.11, 5.30) | 15.30 (14.37, 16.29) | 1.06 (1.02, 1.11) | 0.88 (0.83, 0.94) | 1.29 (1.26, 1.33) | 1.05 (1.00, 1.11) | 0.57 (0.53, 0.62) | 0.87 (0.81, 0.94) | 0.76 (0.62, 0.94) |
| 8-14d | | 53.42 (50.02, 57.06) | 2.48 (2.22, 2.77) | 0.92 (0.58, 1.47) | 2.20 (1.38, 3.51) | 14.38 (13.33, 15.50) | 1.23 (1.16, 1.31) | 0.97 (0.89, 1.06) | 1.71 (1.66, 1.75) | 1.45 (1.37, 1.54) | 0.58 (0.53, 0.65) | 0.88 (0.80, 0.98) | 0.58 (0.42, 0.80) |
| 15-28d | | 51.43 (48.14, 54.95) | 2.91 (2.64, 3.22) | 1.13 (0.74, 1.71) | 2.22 (1.41, 3.50) | 13.92 (12.91, 15.01) | 1.18 (1.12, 1.26) | 1.06 (0.98, 1.15) | 1.58 (1.53, 1.63) | 2.02 (1.93, 2.12) | 0.76 (0.70, 0.83) | 1.05 (0.96, 1.15) | 0.75 (0.57, 0.99) |
| 29-60d | | 50.20 (46.97, 53.66) | 4.03 (3.70, 4.38) | 0.98 (0.63, 1.53) | 4.55 (3.27, 6.32) | 14.47 (13.44, 15.59) | 1.32 (1.25, 1.39) | 1.10 (1.02, 1.19) | 1.47 (1.42, 1.52) | 2.41 (2.31, 2.50) | 0.92 (0.85, 0.99) | 1.49 (1.39, 1.61) | 1.46 (1.19, 1.78) |
| 61-90d | | 48.87 (45.40, 52.60) | 5.86 (5.31, 6.46) | 1.32 (0.75, 2.33) | 4.72 (2.97, 7.52) | 15.33 (13.95, 16.85) | 1.53 (1.42, 1.64) | 1.32 (1.19, 1.47) | 1.44 (1.37, 1.51) | 2.26 (2.13, 2.41) | 1.22 (1.11, 1.34) | 1.83 (1.66, 2.01) | 1.92 (1.48, 2.48) |
| >90d | | 46.89 (43.83, 50.17) | 6.75 (6.32, 7.20) | 2.23 (1.65, 3.01) | 5.38 (3.95, 7.32) | 17.68 (16.50, 18.94) | 1.73 (1.65, 1.81) | 1.60 (1.51, 1.71) | 1.33 (1.28, 1.38) | 2.20 (2.10, 2.30) | 1.43 (1.35, 1.52) | 2.12 (1.99, 2.25) | 2.44 (2.09, 2.86) |
| **Manual Therapy (MT)** | | | | | | | | | | | | | |
| 0-7d | 11.82 (11.53, 12.11) | | 2.04 (1.85, 2.25) | 1.29 (0.92, 1.81) | 6.92 (5.24, 9.15) | 10.88 (10.27, 11.53) | 0.99 (0.94, 1.05) | 0.91 (0.84, 0.99) | 1.18 (1.14, 1.22) | 0.98 (0.92, 1.05) | 0.56 (0.51, 0.62) | 0.91 (0.83, 1.00) | 0.64 (0.48, 0.84) |
| 8-14d | 12.20 (11.91, 12.51) | | 1.72 (1.49, 1.98) | 1.08 (0.66, 1.77) | 4.85 (3.19, 7.37) | 9.31 (8.62, 10.05) | 1.18 (1.10, 1.27) | 0.93 (0.83, 1.03) | 1.62 (1.56, 1.67) | 1.35 (1.26, 1.45) | 0.56 (0.49, 0.64) | 0.91 (0.80, 1.02) | 0.76 (0.54, 1.07) |
| 15-28d | 12.03 (11.74, 12.33) | | 2.07 (1.84, 2.34) | 1.26 (0.83, 1.93) | 4.11 (2.70, 6.25) | 9.93 (9.26, 10.64) | 1.16 (1.08, 1.24) | 1.06 (0.97, 1.16) | 1.54 (1.49, 1.60) | 1.90 (1.80, 2.00) | 0.75 (0.68, 0.83) | 1.06 (0.96, 1.18) | 0.68 (0.49, 0.95) |
| 29-60d | 11.83 (11.53, 12.13) | | 2.88 (2.60, 3.18) | 0.68 (0.39, 1.20) | 7.45 (5.40, 10.28) | 9.75 (9.09, 10.46) | 1.33 (1.25, 1.41) | 1.04 (0.95, 1.14) | 1.45 (1.40, 1.50) | 2.29 (2.19, 2.40) | 0.84 (0.76, 0.92) | 1.49 (1.37, 1.61) | 1.16 (0.90, 1.49) |
| 61-90d | 11.17 (10.81, 11.53) | | 4.25 (3.78, 4.77) | 0.89 (0.42, 1.87) | 8.35 (5.41, 12.89) | 10.66 (9.73, 11.68) | 1.47 (1.35, 1.59) | 1.19 (1.05, 1.35) | 1.41 (1.33, 1.49) | 2.21 (2.06, 2.37) | 1.24 (1.11, 1.38) | 1.91 (1.72, 2.12) | 1.77 (1.31, 2.40) |
| >90d | 11.10 (10.80, 11.41) | | 4.86 (4.50, 5.24) | 1.93 (1.36, 2.73) | 10.14 (7.61, 13.51) | 12.09 (11.35, 12.87) | 1.76 (1.67, 1.85) | 1.66 (1.55, 1.79) | 1.32 (1.27, 1.37) | 2.22 (2.12, 2.33) | 1.43 (1.34, 1.53) | 2.25 (2.11, 2.40) | 2.35 (1.96, 2.81) |
| **Manipulation - Chiropractic (CMT)** | | | | | | | | | | | | | |
| 0-7d | 2.86 (2.69, 3.04) | 2.38 (2.16, 2.61) | | 0.87 (0.42, 1.83) | 3.35 (1.93, 5.81) | 8.62 (7.99, 9.31) | 0.96 (0.86, 1.07) | 1.01 (0.88, 1.16) | 0.88 (0.81, 0.96) | 0.70 (0.61, 0.81) | 0.83 (0.72, 0.96) | 0.90 (0.77, 1.06) | 0.51 (0.29, 0.90) |
| 8-14d | 2.68 (2.44, 2.95) | 2.47 (2.17, 2.82) | | 1.29 (0.54, 3.10) | 4.27 (2.13, 8.55) | 8.80 (7.93, 9.77) | 1.15 (1.00, 1.33) | 1.26 (1.05, 1.50) | 0.89 (0.79, 1.00) | 0.81 (0.67, 0.97) | 1.02 (0.85, 1.22) | 0.69 (0.52, 0.90) | 0.44 (0.18, 1.05) |
| 15-28d | 2.47 (2.27, 2.68) | 2.32 (2.07, 2.59) | | 0.85 (0.36, 2.05) | 3.53 (1.89, 6.60) | 8.42 (7.70, 9.21) | 1.10 (0.97, 1.23) | 1.07 (0.91, 1.25) | 0.94 (0.86, 1.03) | 0.95 (0.82, 1.09) | 1.09 (0.95, 1.26) | 0.94 (0.78, 1.13) | 0.47 (0.23, 0.93) |
| 29-60d | 2.54 (2.38, 2.71) | 2.28 (2.08, 2.50) | | 0.98 (0.51, 1.89) | 4.05 (2.53, 6.48) | 8.49 (7.89, 9.14) | 1.17 (1.07, 1.29) | 1.02 (0.89, 1.16) | 0.83 (0.77, 0.90) | 0.78 (0.69, 0.89) | 1.27 (1.15, 1.41) | 0.80 (0.68, 0.94) | 0.67 (0.42, 1.06) |
| 61-90d | 2.61 (2.43, 2.82) | 2.57 (2.33, 2.84) | | 0.62 (0.23, 1.66) | 3.22 (1.72, 6.01) | 8.04 (7.35, 8.78) | 1.31 (1.18, 1.45) | 1.04 (0.89, 1.21) | 0.92 (0.84, 1.00) | 0.92 (0.80, 1.05) | 1.23 (1.08, 1.39) | 0.93 (0.78, 1.11) | 0.58 (0.32, 1.05) |
| >90d | 2.84 (2.72, 2.97) | 2.57 (2.41, 2.74) | | 1.20 (0.77, 1.86) | 5.44 (3.99, 7.43) | 9.58 (9.08, 10.10) | 1.45 (1.37, 1.54) | 1.28 (1.17, 1.39) | 1.07 (1.02, 1.12) | 1.11 (1.03, 1.20) | 1.27 (1.18, 1.37) | 1.37 (1.25, 1.49) | 1.47 (1.17, 1.85) |
| **Manipulation - Osteopathic (OMT)** | | | | | | | | | | | | | |
| 0-7d | 1.36 (1.18, 1.58) | 0.72 (0.55, 0.94) | 0.71 (0.47, 1.05) | | 1.81 (0.68, 4.83) | 1.63 (1.29, 2.07) | 0.41 (0.32, 0.53) | 0.48 (0.36, 0.65) | 0.36 (0.29, 0.45) | 0.30 (0.21, 0.41) | 0.30 (0.21, 0.43) | 0.50 (0.36, 0.69) | 0.35 (0.13, 0.94) |
| 8-14d | 1.60 (0.93, 2.75) | 1.51 (0.74, 3.08) | insufficient data | | insufficient data | 2.81 (1.38, 5.73) | 0.97 (0.51, 1.85) | 1.06 (0.48, 2.36) | 0.83 (0.50, 1.38) | 0.62 (0.25, 1.54) | 0.73 (0.29, 1.82) | 1.72 (0.90, 3.28) | 3.05 (0.80, 11.64) |
| 15-28d | 2.17 (1.50, 3.15) | 2.31 (1.44, 3.68) | 1.21 (0.41, 3.59) | | insufficient data | 2.15 (1.03, 4.47) | 0.95 (0.54, 1.68) | 1.46 (0.82, 2.58) | 0.70 (0.42, 1.15) | 0.83 (0.42, 1.62) | 1.25 (0.71, 2.22) | 1.13 (0.54, 2.35) | 1.17 (0.17, 8.06) |
| 29-60d | 1.81 (1.27, 2.58) | 1.67 (1.03, 2.69) | 1.34 (0.58, 3.11) | | 11.88 (3.93, 35.90) | 2.62 (1.54, 4.46) | 0.63 (0.35, 1.16) | 1.19 (0.70, 2.02) | 0.76 (0.52, 1.12) | 1.03 (0.64, 1.65) | 1.30 (0.83, 2.05) | 1.88 (1.22, 2.90) | 1.55 (0.40, 6.07) |
| 61-90d | 1.44 (0.77, 2.71) | 1.75 (0.87, 3.52) | 0.61 (0.09, 4.18) | | insufficient data | 2.72 (1.24, 5.95) | 0.96 (0.48, 1.94) | 1.48 (0.74, 2.97) | 0.87 (0.52, 1.47) | 0.90 (0.41, 1.97) | 0.85 (0.35, 2.08) | 2.28 (1.29, 4.04) | 1.77 (0.26, 12.09) |
| >90d | 2.30 (1.88, 2.81) | 2.39 (1.85, 3.10) | 2.32 (1.51, 3.55) | | 15.18 (7.72, 29.84) | 2.85 (2.01, 4.05) | 1.32 (1.02, 1.70) | 1.65 (1.23, 2.23) | 0.77 (0.59, 1.00) | 1.29 (0.97, 1.71) | 1.47 (1.10, 1.96) | 1.74 (1.27, 2.39) | insufficient data |
| **Acupuncture (Acu)** | | | | | | | | | | | | | |
| 0-7d | 2.81 (2.35, 3.37) | 3.95 (3.29, 4.74) | 1.84 (1.08, 3.13) | 5.19 (1.98, 13.59) | | 7.26 (6.02, 8.76) | 0.88 (0.61, 1.27) | 0.24 (0.09, 0.64) | 0.53 (0.36, 0.76) | 0.67 (0.42, 1.07) | 0.58 (0.33, 1.01) | 0.92 (0.55, 1.53) | 0.44 (0.06, 3.10) |
| 8-14d | 2.58 (1.83, 3.65) | 4.39 (3.29, 5.85) | 2.42 (1.08, 5.42) | 4.09 (0.59, 28.22) | | 5.62 (3.70, 8.55) | 1.13 (0.65, 1.97) | 0.38 (0.10, 1.47) | 0.68 (0.39, 1.18) | 0.99 (0.51, 1.90) | 1.00 (0.48, 2.05) | 1.12 (0.50, 2.50) | 1.39 (0.20, 9.58) |
| 15-28d | 2.04 (1.41, 2.96) | 3.61 (2.66, 4.90) | 5.05 (3.28, 7.77) | 3.05 (0.44, 21.17) | | 6.76 (4.97, 9.19) | 1.03 (0.62, 1.72) | 0.29 (0.07, 1.11) | 0.62 (0.37, 1.03) | 1.05 (0.61, 1.81) | 0.99 (0.53, 1.85) | 1.16 (0.59, 2.29) | 1.04 (0.15, 7.18) |
| 29-60d | 1.99 (1.48, 2.68) | 3.27 (2.52, 4.26) | 4.43 (3.06, 6.42) | 3.74 (0.95, 14.70) | | 6.13 (4.71, 7.98) | 0.98 (0.65, 1.48) | 0.79 (0.43, 1.46) | 0.69 (0.48, 1.00) | 1.36 (0.95, 1.94) | 1.14 (0.73, 1.78) | 1.23 (0.73, 2.05) | 1.27 (0.32, 4.99) |
| 61-90d | 2.80 (2.16, 3.63) | 4.31 (3.39, 5.48) | 3.88 (2.37, 6.33) | insufficient data | | 6.06 (4.39, 8.35) | 1.01 (0.62, 1.65) | 1.41 (0.84, 2.37) | 1.11 (0.82, 1.51) | 2.07 (1.52, 2.82) | 1.66 (1.09, 2.53) | 2.53 (1.73, 3.71) | 1.85 (0.48, 7.21) |
| >90d | 2.48 (2.11, 2.93) | 3.88 (3.34, 4.51) | 3.98 (3.04, 5.20) | 5.88 (2.84, 12.17) | | 7.36 (6.35, 8.55) | 1.55 (1.27, 1.89) | 1.03 (0.72, 1.46) | 0.82 (0.66, 1.02) | 1.48 (1.18, 1.85) | 1.16 (0.86, 1.56) | 1.88 (1.44, 2.45) | 2.00 (0.97, 4.12) |

Cells in red are not different than the reference of if the specific service was not performed (p=.05)
