## Supplement - State Summary for "Low back pain service utilization and costs: association with timing of first-line services for individuals initially contacting a physician specialist. A retrospective cohort study"

### Supplement - Episode count by home address state of individual with low back pain

| State | Episodes | % |
|---|---|---|
| Total | 98992 | 100.0% |
| FL | 13332 | 13.5% |
| TX | 12582 | 12.7% |
| NY | 5118 | 5.2% |
| CA | 4520 | 4.6% |
| GA | 4301 | 4.3% |
| MD | 4060 | 4.1% |
| IL | 3966 | 4.0% |
| NC | 3949 | 4.0% |
| OH | 3822 | 3.9% |
| VA | 3268 | 3.3% |
| CO | 3248 | 3.3% |
| MO | 2956 | 3.0% |
| LA | 2887 | 2.9% |
| AZ | 2846 | 2.9% |
| WI | 2637 | 2.7% |
| NJ | 2284 | 2.3% |
| MN | 2156 | 2.2% |
| TN | 1972 | 2.0% |
| IN | 1930 | 2.0% |
| PA | 1796 | 1.8% |
| CT | 1343 | 1.4% |
| NE | 995 | 1.0% |
| OK | 968 | 1.0% |
| MS | 923 | 0.9% |
| MI | 915 | 0.9% |
| UT | 877 | 0.9% |
| MA | 810 | 0.8% |

| State | Episodes | % |
|---|---|---|
| WA | 790 | 0.8% |
| SC | 758 | 0.8% |
| AR | 749 | 0.8% |
| KY | 676 | 0.7% |
| IA | 620 | 0.6% |
| AL | 569 | 0.6% |
| KS | 565 | 0.6% |
| RI | 524 | 0.5% |
| NV | 482 | 0.5% |
| DC | 479 | 0.5% |
| OR | 407 | 0.4% |
| NM | 199 | 0.2% |
| WV | 176 | 0.2% |
| NH | 163 | 0.2% |
| DE | 115 | 0.1% |
| ID | 91 | 0.1% |
| ND | 85 | 0.1% |
| VI | 84 | 0.1% |
| WY | 72 | 0.1% |
| ME | 64 | 0.1% |
| SD | 41 | 0.0% |
| MT | 34 | 0.0% |
| HI | 16 | 0.0% |
| AK | 10 | 0.0% |
| PR | 8 | 0.0% |
| VT | 7 | 0.0% |
| Unknown | 747 | 0.8% |
